# Regional, circuit, and network heterogeneity of brain abnormalities in psychiatric disorders

**DOI:** 10.1101/2022.03.07.22271986

**Authors:** Ashlea Segal, Linden Parkes, Kevin Aquino, Seyed Mostafa Kia, Thomas Wolfers, Barbara Franke, Martine Hoogman, Christian F. Beckmann, Lars T. Westlye, Ole A. Andreassen, Andrew Zalesky, Ben J. Harrison, Christopher Davey, Carles Soriano-Mas, Narcís Cardoner, Jeggan Tiego, Murat Yücel, Leah Braganza, Chao Suo, Michael Berk, Sue Cotton, Mark A. Bellgrove, Andre F. Marquand, Alex Fornito

## Abstract

The substantial individual heterogeneity that characterizes mental illness is often ignored by classical case-control designs that rely on group mean comparisons. Here, we present a comprehensive, multiscale characterization of individual heterogeneity of brain changes in 1294 cases diagnosed with one of six conditions and 1465 matched healthy controls. Normative models identified that person-specific deviations from population expectations for regional grey matter volume were highly heterogeneous, affecting the same area in <7% of people with the same diagnosis. However, these deviations were embedded within common functional circuits and networks in up to 56% of cases. The salience/ventral attention system was implicated transdiagnostically, with other systems selectively involved in depression, bipolar disorder, schizophrenia, and ADHD. Our findings indicate that while phenotypic differences between cases assigned the same diagnosis may arise from heterogeneity in the location of regional deviations, phenotypic similarities are attributable to dysfunction of common functional circuits and networks.

## INTRODUCTION

The neurobiological mechanisms of mental illness are elusive. Thousands of neuroimaging studies have documented a diverse array of structural and functional brain changes associated with specific psychiatric diagnoses and meta-analyses of this work have identified the brain regions that are most consistently affected in each condition, revealing both diagnosis-specific and transdiagnostic effects^1,2,11–15,3–10^. However, despite this substantial research effort, pathophysiological processes are poorly understood, and clinically useful biomarkers are lacking.

One reason for this limited progress may be a continued reliance on case-control designs, which focus on comparison of group-mean values and ignore the considerable clinical heterogeneity often shown by individuals with the same diagnosis^16–19^. Indeed, recent neuroimaging studies that investigate individual-specific patterns of brain deviations have found that group average differences are not likely to be representative of individual cases^20–26^. These individual-specific inferences are often performed using normative modelling^27^, which involves training a model to learn normative expectations for a brain phenotype, such as grey matter volume (GMV), given an individual’s age, sex, or other relevant characteristics. The model predictions can then be used to define a normative range of variation against which new individuals are compared. Fitting the model to data for multiple brain regions allows one to obtain a personalized deviation map that quantifies the extent to which the individual deviates from population norms within each brain region, thus enabling the identification of brain regions associated with unusually small or large phenotypic values, termed extreme deviations, in an individual. Normative modelling studies of diverse MRI-derived phenotypes in attention-deficit/hyperactivity disorder^20^ (ADHD), bipolar disorder (BP), schizophrenia^21,24,26^ (SCZ), autism spectrum disorder^22,23,25^ (ASD) have found that, while cases often have more extreme deviations than healthy controls, the specific location of these deviations varies considerably between individuals with the same diagnosis.

This extreme regional heterogeneity of individual brain deviations aligns with the well-described clinical heterogeneity often associated with specific psychiatric diagnoses^16–19^, but raises an important question: if cases show little overlap in the anatomical locations of their GMV deviations, what then explains phenotypic similarities between people assigned the same diagnostic label? It seems reasonable to assume that such similarities are driven by some common aspect of neural dysfunction across individuals, but the findings of normative modelling studies suggest otherwise. One possible explanation is that while the specific location of brain deviations may vary across individuals, these deviations aggregate within common circuits or neural systems. The brain is a connected network, and pathological processes often affect distributed, interconnected systems^28–30^, meaning that it is possible for deviations in disparate loci to impact the function of common, connected areas. This principle has been demonstrated through lesion network mapping studies of neurological patients sharing a common motor, perceptual, or cognitive syndrome^31,32^. Specifically, this work has shown that patients often show little overlap in the anatomical location of their lesions, but that lesioned sites are often functionally coupled to common areas^31,33,42–51,34–41^. The clinical expression of such syndromes thus appears to be more closely related to dysfunction of these coupled areas rather than of the lesioned region itself.

Here, we considered whether a similar process is at play in psychiatric disorders. Specifically, we investigated whether anatomically heterogeneous regional brain deviations within psychiatric disorders are functionally coupled to common areas and networks. To this end, we developed a new framework to integrate normative models of GMV with elements of lesion network mapping to map the functional circuits and extended networks within which regional GMV deviations are embedded. We used this approach to derive a multiscale characterization of neural heterogeneity across 1294 individuals diagnosed with one of six disorders: ADHD, ASD, BP, major depressive disorder (MDD), obsessive-compulsive disorder (OCD), and SCZ. Inspired by studies of patients with brain lesions^31,32^, we tested the hypothesis that anatomically heterogeneous regional GMV deviations in each disorder are functionally coupled with common sites, either within a functional circuit or an extended functional network (see Figure 1 for a schematic explanation). Our transdiagnostic, multiscale approach allowed us to comprehensively understand the extent of neural heterogeneity within each disorder while also revealing commonalities and differences between disorders.

**Figure 1.**
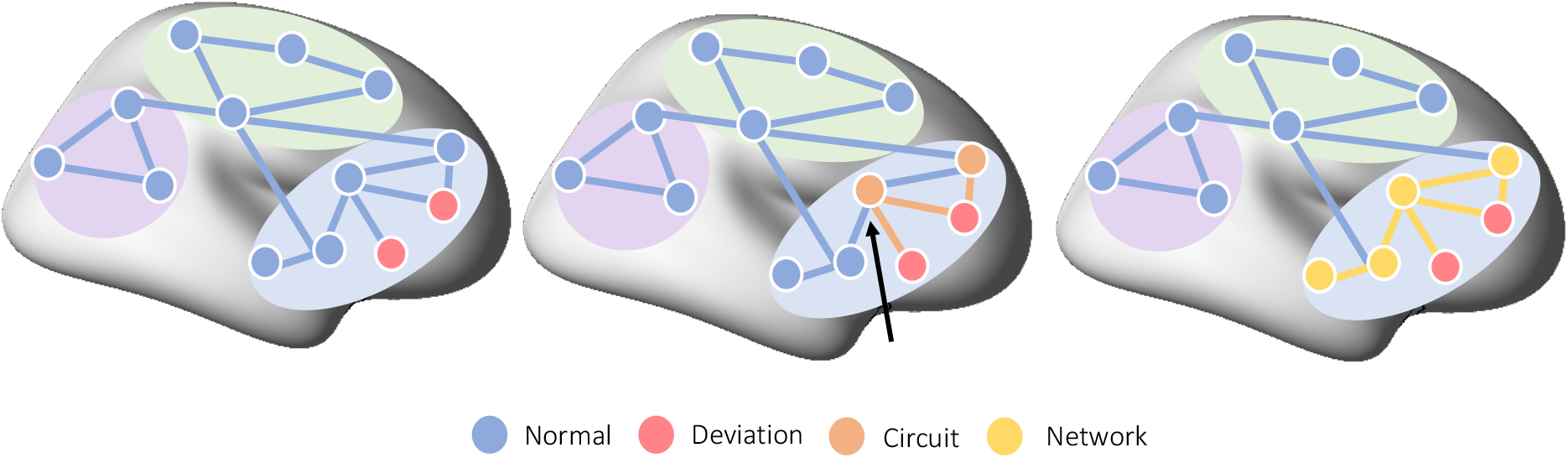
Characterizing neural heterogeneity at the level of brain regions, functional circuits, and extended networks. Schematic showing how neural heterogeneity can be characterized at different scales. Nodes represent different brain regions, edges represent functional coupling between nodes, and colored areas correspond to different functional networks of the brain. At the regional level (left), deviations from normative model predictions are localized to specific brain regions in each individual. Red nodes show the locations of such deviations mapped in two different people. A circuit-level analysis (middle) reveals areas the areas that are functionally coupled to the deviant loci. In this work, we define a functional circuit as the set of regions that show significant functional coupling with a specific deviant region (orange). In this example, the two deviant areas are coupled to a common region (black arrow) despite being located in different areas themselves. (c) These circuits can be embedded within extended networks (right) that include regions which may not be directly coupled to the deviant regions, but which nonetheless participate within the same functional system (yellow).

## RESULTS

### Sample Characteristics

We examined data for 1465 healthy controls (HC; 54.47% male) and 1294 cases, taken from 14 different studies and 25 different scan sites. The clinical sample comprised 202 individuals with ASD (100% male), 153 individuals with ADHD (41.18% male), 228 individuals with BP (47.37% male), 161 individuals with MDD (34.16% male), 167 individuals with OCD (50.30% male), and 383 participants with SCZ (62.14% male). The scanner details, sample size, and demographic characteristics of each scan site, after various exclusions based on data quality and other criteria (see Methods), are presented in Table S1 (for age distributions, see Figure S1).

### Normative Modelling

We used an established pipeline to obtain voxel-wise estimates of GMV in each participant^52,53^ (see Methods). Voxel-wise GMV estimates were converted into regional estimates for 1032 brain areas defined according to previously validated, functionally-constrained parcellations^54,55^ (1000 cortical regions, 32 subcortical regions; Figure 2a). We then defined a training set comprising 1196 HCs (55.02% male ; Figure 2a; HC_train_) to train, separately in each region, a normative model based on hierarchical Bayesian regression^56^ (HBR), thus establishing a normative GMV range given an individual’s age, sex, and scan site (Figure 2b). HC_train_ spanned the age range of cases, enabling predictions for ages between 18 and 64 years (Figure S2). The remaining 269 controls (52.04% male; age range 18-62) were held-out as a test set (HC_test_) to establish a normative benchmark for comparison with each clinical group (Figure 2c). A detailed explanation of our modelling procedure is provided in the Methods. Model fit statistics and checks for residual site effects are provided in Figure S3 and Table S2.

**Figure 2.**
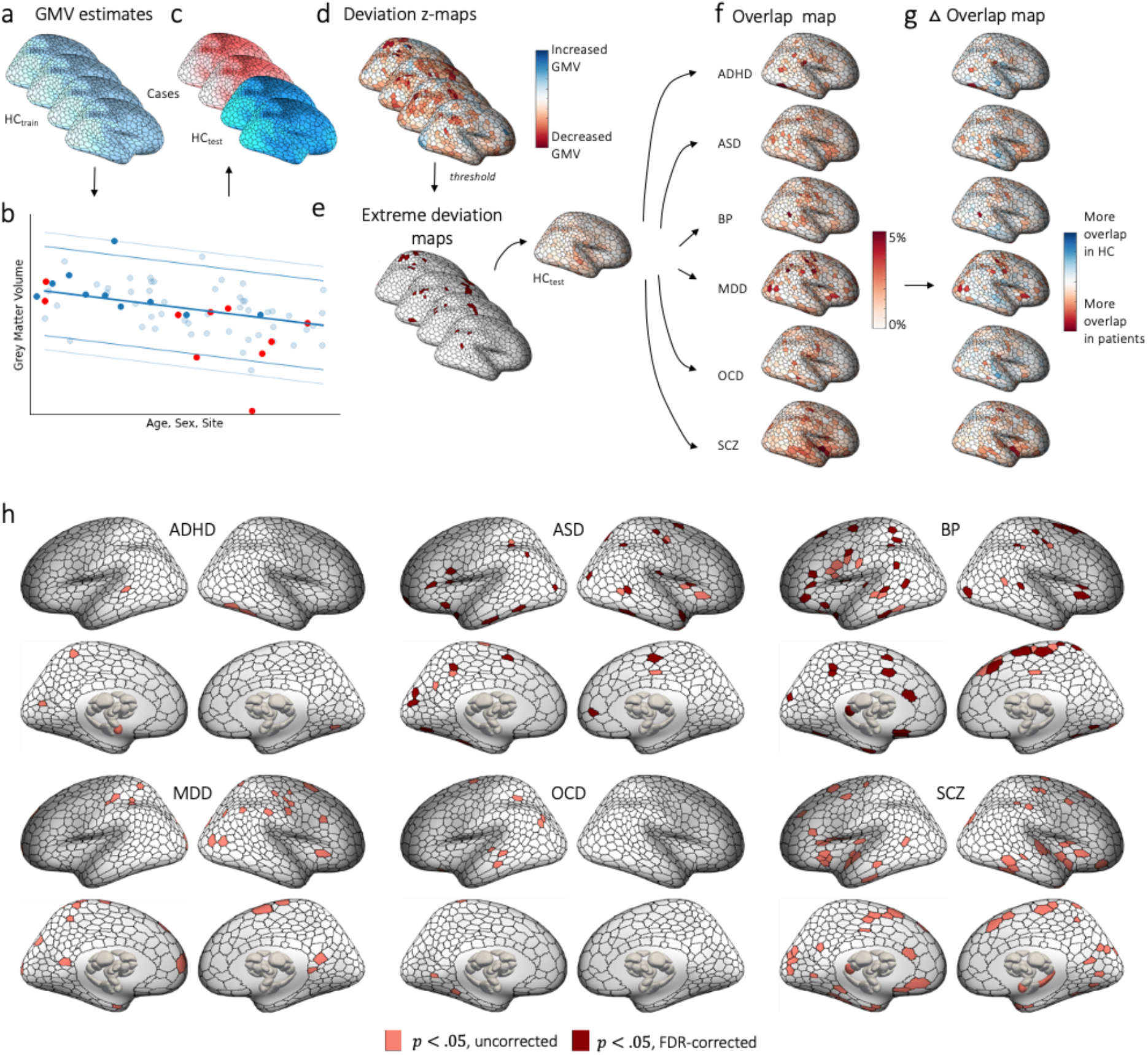
Regional heterogeneity of extreme negative GMV deviations in each disorder. (a-f) Workflow for characterizing regional-level heterogeneity. (a) GMV maps for each individual were parcellated into 1000 cortical and 32 subcortical regions. (b) The training dataset, HC_train_, was used to train a normative model to make predictions about regional GMV values given an individual’s age, sex, and scan site. (c) The predictions for held-out controls (HC_test_) and cases were then compared to empirical GMV estimates. Model predictions for one region, showing individuals in the training set (HC_train_; light blue) and the held-out control (HC_test_; dark blue) and clinical groups (red). Solid and dashed lines indicate the 99^th^ and 95^th^ centiles, respectively. (d) For each individual, deviations from model predictions were quantified as a deviation z-map. (e) This deviation map was then thresholded at z > |2.6| to identify extreme deviations. (f) For the HC_test_ and each clinical group, we quantified the proportion of individuals showing an extreme deviation in a given brain region, yielding an extreme deviation overlap map. (g) We subtracted the HC_test_ overlap map from each clinical group’s overlap map to obtain an overlap difference map (Δ overlap map) for each clinical group and then evaluated the magnitude of this difference (see Figure S4a-d for details). (h) Shows the cortical and subcortical surface renderings showing regions with significantly greater overlap of extreme negative GMV deviations in cases compared to controls.

For each individual in the clinical and HC_test_ groups, we quantified the degree to which regional GMV estimates deviated from normative model predictions as a *z*-score (termed *Deviation z-maps*; see Methods and Figure 2d). Following Wolfers et al.^21^, a z-score > |2.6| defined an extreme deviation (Figure 2e), corresponding to p < .005, uncorrected (see Methods for a discussion of thresholding issues). Figure S4 shows the distribution of positive and negative person-specific deviation burden scores (i.e., the total number of extreme deviations identified in a person) across individuals, stratified by diagnostic group. Nonparametric rank sum tests indicated that people with BP, MDD, OCD and SCZ showed a higher burden of extreme negative deviations compared to HC_test_(*p*<0.003), indicating lower GMV than age- and sex-specific expectations. Only people with ASD showed a higher positive deviation burden than controls (*p*<0.001, descriptive statistics provided in Table S3). Critically, rank correlations indicated scan quality (as measured by CAT12’s IQR) was not meaningfully associated with extreme deviation burden (HC_test_: *rho* = 0.04, *p*=0.51; cases: *rho* = 0.03, *p*=0.34). In the following, we focus on a multiscale characterization of negative GMV deviations (i.e., areas where volume is lower than normative expectations), as these have been most extensively studied in the psychiatric literature. Results for our analysis of positive GMV deviations can be found in Figures S17-24 and Table S4-5, and a discussion is in the Supplementary Material (*Discussion of positive GMV deviations*).

### Characterizing neural heterogeneity at the level of brain regions

We characterized regional heterogeneity in GMV (Figure 1 left) by assessing the degree of spatial overlap in the location of each individual’s extreme negative deviations (Z < -2.6, i.e., lower GMV than expected). Specifically, we quantified the proportion of individuals showing an extreme deviation in each parcellated brain area separately for each diagnostic group and the HC_test_ cohort (Figure 2f). Across all groups, the majority of individual showed at least on extreme deviation, ranging from 75.82% in ADHD to 88.51% in SCZ. Nonetheless, across all groups and all 1032 brain regions, the maximum percentage overlap never exceeded 7% (ADHD: 3.27%, ASD: 4.95%, BP: 5.26%, MDD: 6.21%, OCD: 4.19%, SCZ: 4.96% HC: 2.97%). Hence, while individual extreme deviations were common, they were rarely found in consistent locations across individuals with the same diagnosis (Figure S5).

We next compared the degree of spatial overlap between each clinical group and controls by subtracting their respective percentage overlap values in each region (Figure 2g). The statistical significance of the observed Δ overlap values in each region was assessed with respect to an empirical null distribution generated by shuffling group labels (see Methods and Figure S6a-d). While each disorder showed isolated areas of greater overlap at uncorrected levels, only ASD (32 regions) and BD (45 regions) showed differences that survived False Discovery Rate (FDR) correction (*p*_*FDR*_<0.05, two-tailed; Figure 2h). These differences were scattered throughout the cortex and rarely aggregated into spatially structured clusters. Few regions showed significantly greater overlap in controls compared to cases (Figure S7). Repeating the analyses using a method that integrates across a range of thresholds rather than relying on a single threshold for defining extreme deviations^57^ yielded similar findings (see Methods; Figure S8-S10). Collectively, these results extend past findings^20,21,26^ to indicate that minimal spatial overlap in the location of person-specific extreme GMV deviations is a general characteristic of psychiatric illness.

### Characterizing neural heterogeneity at the level of functional circuits

We next asked whether the regionally heterogeneous extreme deviations identified in each clinical group show significant functional coupling (FC) with common, remote areas, thus yielding greater inter-individual consistency at the level of functionally coupled circuitry (Figure 1, middle). To this end, we took each region showing an extreme deviation in each participant (Figure 3a) and mapped its pattern of whole-brain FC in an independent sample of 150 unrelated healthy controls (HCP_150_) to establish the normative pattern of expected FC for the deviant region (Figure 3b). We thresholded (*p*_*FWE*_ < 0.025) and binarized each deviant-related FC map (Figure 3c), and took the union of these thresholded maps across all extreme deviations for a given person (Figure 3d; for a discussion of thresholding issues, see Discussion), yielding a map that represents all areas showing significant FC with at least one extreme deviation in that individual. Next, we estimated, for each region, the proportion of individuals within each group for whom that region showed significant FC with an extreme deviation (Figure 3e). This analysis revealed that the overlap observed in sites functionally coupled to deviant regions was much higher, in absolute terms, than the overlap observed in the locations of the extreme deviations themselves (Figure S11). For instance, the maximum circuit-level overlap observed across regions was 33% in the HC_test_ group and ranged between 39% (ADHD) and 53% (SCZ) in the clinical groups.

**Figure 3.**
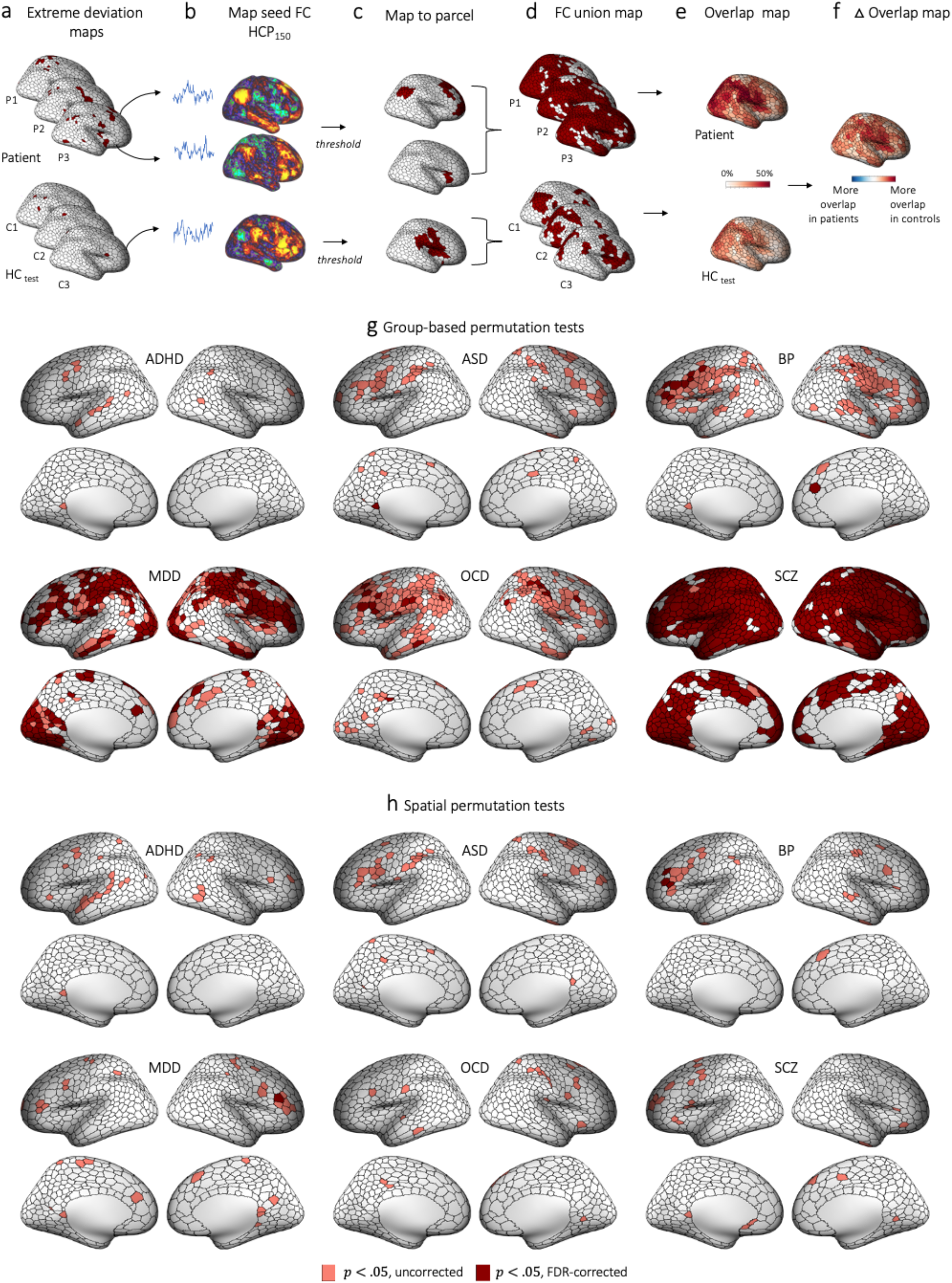
Functional circuit heterogeneity of extreme negative GMV deviations in each disorder. (a-f) Workflow for characterizing circuit-level heterogeneity. (a) For each participant in the HC_test_ and each clinical group, we took each brain region showing an extreme deviation. (b) For each individual in an independent sample of controls (HC_150_), we extracted a representative time course from each deviant region and mapped the areas to which it is functionally coupled using a seed-related FC analysis. Shown here are two FC maps for two different deviant loci identified in one individual. (c) We thresholded and binarized each FC map associated with a given extreme deviation. Note, that no subcortical regions survived this thresholding procedure. (d) We took the union of the thresholded maps across all deviant FC maps to obtain a single map of all areas showing direct FC with one or more deviant areas for a given individual. (e) For the HC_test_ and each clinical group, we quantified the proportion of individuals showing significant FC in a given region, yielding an extreme deviation FC overlap map. (f) We subtracted the HC_test_ FC overlap map from each clinical group’s FC overlap map to obtain an FC Δ overlap map for each clinical group. (g) Group differences in circuit-level overlap were evaluated with respect to two empirical null models (see Figure S6 for details). (g) and (h) show cortical surface renderings of regions with significantly greater overlap in cases compared to controls in areas functionally coupled to extreme deviations identified using group-based or spatial permutation tests, respectively.

To a certain extent, one should expect overlap to be higher at the level of FC union maps (Figure 3d-e) than regional deviation maps (Figure 2e-f), since any given deviant area can show FC with multiple other regions, increasing the likelihood that common areas will be implicated across individuals. For this reason, regional circuit-level overlap maps in each clinical group must be contrasted with the HC_test_ overlap map, which provides a normative benchmark for the expected level of overlap in FC union values (Figure 3f). A critical consideration in these contrasts concerns the effect of total deviation burden, which differs between groups (e.g., Figure S4). For instance, the total number of extreme deviations identified in individuals diagnosed with SCZ was 4951 compared to only 1410 in HC_test_. This discrepancy means that more deviant-related FC maps will be used in the FC union maps of SCZ individuals, thereby increasing the chance of observing higher overlap across individuals. On the one hand, this higher circuit-level overlap will have real phenotypic consequences, since a higher deviation burden is an intrinsic and expected characteristic of psychiatric disorders and these deviations are likely to impact circuit-level function in a way that affects behavior. On the other hand, it is important to know whether the overlap is driven simply by group differences in deviation burden or reflects a preferential targeting of the circuit by the disorder in question. We therefore evaluated the statistical significance of regional group differences in circuit-level overlap using two different permutation tests designed to disentangle these effects (see Methods and Figure S6 for details). The first, *group-based permutation test*, relied on shuffling the group labels of the individual-specific FC union maps (Figure S6a-d) and assessed differences in overall circuit-level overlap, regardless of group differences in total deviation burden, thereby characterizing group differences in their “natural” state. The second, *spatial permutation test*, evaluated group differences with respect to a null distribution that preserves the number of deviation-related FC maps contributed by each individual, thus matching differences in the total deviation burden of each group (see Methods and Figure S6e-k). This analysis therefore tests whether observed group differences in circuit-level overlap are greater than expected for FC maps generated from the same number of randomly chosen seeds, with the random seeds being selected from a deviation map with the same underlying spatial autocorrelation as the empirical deviation maps (see Methods for further details and interpretation).

Using the group-based permutation tests, we observed significantly greater overlap across wide swathes of cortex for both people with SCZ and MDD (*p*_*FDR*_ < 0.05, two-tailed) compared to controls (Figure 3g). Regions with significantly greater overlap in SCZ were distributed diffusely and included ∼75% of cortical areas. In MDD, ∼31% of areas were implicated, being predominately localized to regions of visual, parietal, somatomotor, frontal, and insula cortices. A similar spatial pattern of findings was observed for people with ASD, OCD, and BP when considering uncorrected results, although fewer regions survived FDR correction. Very few areas showed greater overlap in deviant-related FC in controls compared to cases (Figure S12a). An alternative method for mapping FC results to parcellated regions yielded comparable findings (see Methods and Figures S13, S14a-15a). These findings support the hypothesis that GMV deviations in psychiatric disorders are part of common functional circuits despite being located in anatomically heterogeneous areas.

We next used the spatial permutation test to determine the degree to which the above differences in circuit-level overlap were attributable to deviation burden. The results, shown in Figure 3h, indicate that all disorders show some evidence for greater overlap in regions of left inferior and middle frontal gyri at uncorrected thresholds. However, only three areas in BD and one in MDD survived FDR correction. Similarly, few results survived FDR correction when considering regions showing greater overlap in controls (Figure S12b) and an alternative method for mapping FC results to our regional parcellation yielded comparable findings (see Methods, Figure S14b-15b). Thus, while these findings offer preliminary evidence for the preferential involvement of neural circuitry involving the lateral prefrontal cortex in each disorder, they indicate that the dominant factor explaining greater circuit-level overlap in cases is total deviation burden. In other words, cases show greater overlap at the circuit level largely because they are more likely to express extreme GMV deviations, which in turn increases the probability with which sites functionally coupled to deviant areas will be implicated.

### Characterizing neural heterogeneity at the level of extended functional networks

Our analysis thus far indicates that the loci of regional GMV deviations show marked individual heterogeneity, that cases show substantially greater overlap when considering the functional circuitry of these deviant loci, and that this overlap is largely driven by total deviation burden. However, our circuit-level analysis only focused on areas showing significant FC with deviant regions, and these circuits often form part of larger, extended functional networks that may not be fully mapped by a circuit-level characterization (Figure 1 right). We therefore examined the location of person-specific GMV deviations in relation to canonical functional networks, defined according to a widely-used and validated classification of brain areas into one of 7 cortical networks^54,58^ or 3 subcortical regions^55^, to derive a comprehensive, multiscale characterization of GMV heterogeneity (Figure 4a-c). In this way, if an individual showed an extreme deviation in at least one region affiliated to a given network, the entire network was considered deviant (Figure 4c). Once again, we quantified the proportion of individuals within each group showing a deviation in each network (Figure 4d) and compared these proportions between the HC_test_ and each clinical group (Figure 4e). We then evaluated group differences in network-level overlap using both group-based and spatial permutation tests, as in the circuit-level analysis (Figure S6).

**Figure 4.**
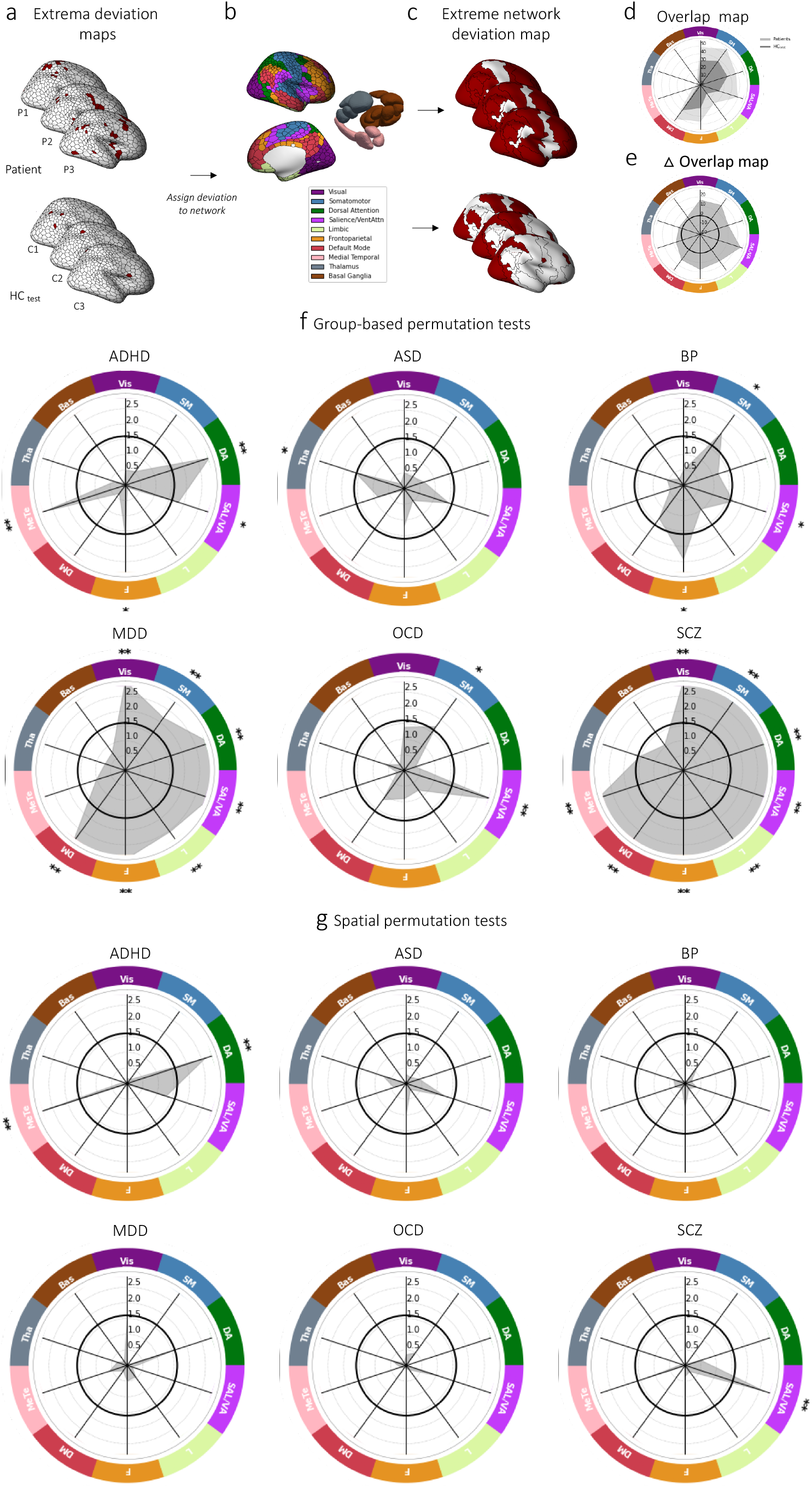
Functional network heterogeneity of extreme negative GMV deviations in each disorder. (a-d) Workflow for characterizing network-level GMV heterogeneity. (a) For each individual in the HC_test_ and each clinical group, we assigned each brain region showing an extreme deviation to one of 10 canonical cortical functional networks or 3 subcortical nuclei (b), such that the entire network was considered deviant if it contained at least one region with an extreme deviation. Panel (c) shows the resulting network-level extreme deviation maps. (d) We quantified the proportion of individuals in each group showing a deviation within each network and compared these proportions to the network overlap in HC_test_. (e) Group differences in network-level overlap were evaluated with respect to two empirical null models (see Figure S6 for details). Panels (f) and (g) show the network-level -log_10_ p-values associated with the difference in percent overlap for extreme negative GMV deviations between each clinical group and the HC_test_ cohort (grey). ** corresponds to *p*_*FDR*_ < 0.05, two-tailed, cases>controls, * corresponds to *p*_*uncorrected*_ < 0.05, two-tailed, cases>controls. The solid black line indicates -log_10_ p = 1.6 (*p*=0.05, two-tailed, uncorrected). Panels (f) and (g) identify networks showing significant differences under group-based or spatial permutation testing, respectively.

The results of these analyses are shown in Figure 4f-d (see Table S4 for a summary of the degree of spatial overlap (%) in each network for each group). Using group-based permutation testing, SCZ and MDD cases showed significantly greater overlap in multiple networks compared to controls (*q* < 0.05, two-tailed, Figure 4f). For SCZ individuals, the differences included all networks except for the thalamus and basal ganglia. In MDD, all cortical networks were implicated. The dorsal attention network and medial temporal lobe were implicated in ADHD and the ventral attention network in OCD. No networks survived multiple comparison correction for ASD and BP. At uncorrected levels, the salience/ventral attention network was implicated across all disorders except for ASD. There were no networks in which controls showed greater overlap (Figure S16a). Using spatial permutation tests with FDR correction, only the salience/ventral attention network showed greater overlap in SCZ, and the medial temporal lobe and dorsal attention networks showed greater overlap in ADHD (Figure 4g). No deviant-related networks showed greater overlap in controls (Figure S16b).

Together, these results align with the circuit-level analysis to indicate that SCZ and MDD are associated with greater network-level overlap in GMV deviations. The effects are more circumscribed in ADHD, ASD, BP, and OCD, although the salience/ventral attention network is implicated across most disorders. These effects are largely driven by total deviation burden, with only SCZ and ADHD showing robust evidence for preferential involvement of the salience/ventral attention and dorsal attention/medial temporal systems, respectively.

## DISCUSSION

We used normative modelling to characterize the heterogeneity of extreme GMV deviations at the level of brain regions, functional circuits, and extended networks in six psychiatric disorders. We showed that heterogeneity between individuals in regional GMV abnormalities is a general feature of psychiatric illness, with relatively few areas showing significantly greater overlap in cases compared to controls and no single area in any disorder showing an extreme deviation in more than 7% of cases across the 1032 regions. However, these heterogeneous loci were often embedded within common functional circuits and networks, typically involving regions of lateral frontal, parietal, and insula cortex. In some cases, >50% of cases with the same diagnosis showed a deviation implicating at least one of these systems. Much of this elevated circuit and network level overlap across individuals was attributable to group differences in total deviation burden rather than the preferential targeting of a specific circuit or network, although we found evidence for selective involvement of prefrontal circuitry in MDD and BD, the salience/ventral attention network in SCZ, and the dorsal attention and medial temporal networks in ADHD. Together, our findings identify a putative neural substrate for both phenotypic differences and similarities between cases with the same diagnosis, with phenotypic differences being attributable to regional heterogeneity in the location of person-specific GMV deviations and phenotypic similarities attributable to the impact of these deviations on common functional circuits and networks.

The high degree of regional heterogeneity in GMV deviations across disorders aligns with past normative modelling studies of GMV in SCZ, ADHD, and BP^20,21,26^. The variable involvement of different regions across individuals may yield distinct clinical phenotypes and therefore drive phenotypic heterogeneity in people with the same diagnosis^16,17,59^. An important avenue of future work will involve precisely characterizing the relationship between GMV deviations and interindividual differences in symptom expression. More generally, the high degree of regional heterogeneity that we observe indicates that group-mean comparisons may not be representative of the specific profile of brain changes apparent in any individual case and may offer an incomplete account of causal pathophysiological mechanisms unless the broader network context in which the differences occur is considered. Multiple mechanisms may explain these circuit- and network-level effects, ranging from transient diaschisis-like effects on distributed circuit/network function through to more prolonged transneuronal degeneration caused by aberrant inter-regional signaling or axonal transport of trophic factors^29^. Identifying the specific causal mechanisms at play in each disorder will be essential for understanding how pathophysiology spreads through connected neural systems in each disorder.

Despite considerable heterogeneity at the regional level, deviations were often coupled to common functional circuits and networks. This higher overlap at circuit and network levels parallels lesion network mapping studies of neurological syndromes, which suggest that many clinical phenotypes are not caused by dysfunction of the lesioned area itself but by its impact on functionally coupled remote areas^32^. Our findings indicate that a similar process may occur in psychiatric disorders, such that anatomically distributed GMV deviations are often coupled within similar circuits and networks, and the consequent dysfunction of these common circuits and networks may drive clinical similarities between people with the same diagnostic label despite extreme heterogeneity in the locations of the deviations themselves. Indeed, our group-based permutation testing indicated that all disorders show some evidence of greater circuit-level overlap than controls. Areas of frontal, parietal, insula, and temporal cortex were implicated across most conditions, and cross-disorder differences were more of degree rather than kind. For instance, differences in circuit-level overlap were spatially circumscribed in ADHD but encompassed nearly the entire brain in SCZ (Figure 3). These findings challenge classical views that specific psychiatric disorders are associated with dysfunction in distinct circuits (e.g.,^60^) and align with evidence that each disorder is associated with complex changes that affect diverse neural systems, often transdiagnostically^61,62^. Accordingly, our network-level analysis revealed greater overlap in the salience/ventral attention network in 5 of the 6 disorders that we considered (Figure 4). The salience/ventral attention network plays a central role in cognitive control^63^, interoceptive awareness^64^, and switching between internally and externally-focused attention^65^. Its dysfunction has been implicated in a diverse range of psychiatric disorders^66–69^, is associated with increased levels of general psychopathology in youth^70^, and shows cross-disorder abnormalities in meta-analyses of classical case-control VBM studies and functional neuroimaging studies^9,71^. Our findings support this past work to suggest that salience/ventral attention network dysfunction may play a critical role in the expression of general psychopathological processes common to diverse diagnoses^72^.

Our spatial permutation test allowed us to evaluate the degree to which circuit and network-level overlap was explained by group differences in total deviation burden. At the circuit level, prefrontal regions were identified as showing greater overlap compared to controls in all clinical groups at *p*_*uncorrected*_ < .05, consistent with extensive evidence for prefrontal dysfunction in each condition ^73–75^. However, only regions of right and left lateral prefrontal cortex (PFC) in MDD and BD, respectively, survived FDR-correction. The right PFC showed 21% overlap in MDD cases, whereas the left PFC showed 14-15% overlap in BP. The lateral PFC is thought to play an important role in cognitive control^76^, with cognitive control deficits commonly reported in both MDD^77^ and BP^78^. Extensive literature has strongly implicated lateral PFC dysfunction in affective disorders generally^79,80^ and in association to disorder-related cognitive control deficits using task-based fMRI studies^73^. Moreover, dorsolateral PFC is typically used as a transcranial magnetic stimulation (TMS) target site to treat depressive symptoms in MDD^81^ and BP^82^ and has been implicated by lesion network mapping as a core site explaining the emergence of depression following stroke^47^. Given that our spatial permutation test did not find any evidence for greater network-level overlap in MDD and BD, our findings point to a strong specificity for circuits coupled to these particular prefrontal areas. Spatial permutation testing also revealed greater network-level overlap for SCZ in the salience/ventral attention network and for ADHD in the dorsal attention network and medial temporal regions. GMV deviations may thus preferentially aggregate within these systems in a disorder-specific way.

In most other cases, we failed to reject the spatial null hypothesis. This result suggests that many case-control differences in circuit/network overlap observed under group-based permutation cannot be attributed to the preferential accumulation of deviations in a particular circuit/network, since the differences are consistent with expectations from comparing the same number of randomly selected seeds in each group. A corollary of this finding is that many GMV deviations in individual cases may emerge at random locations throughout the brain. As these deviations accumulate, they are more likely to be coupled to areas in prefrontal, temporal, parietal and insula cortices, simply because these areas, which are known as connectivity hubs of the brain^83,84^, have a higher probability of being functionally coupled to other regions. Further work investigating the genetic and environmental contributions to individual-specific deviations should help to better understand the causal mechanisms driving their anatomical distribution.

Sample sizes in single site psychiatric neuroimaging studies are often small, so we pooled data from multiple sites to generate a sufficiently large cross-disorder dataset. As a result, the data were collected with different acquisition, recruitment, and clinical assessment protocols. Although we used stringent quality control, and our hierarchical Bayesian model^56^ was able to appropriately parse site-related variance (Table S2), investigating correlations with symptom profiles or other clinically-relevant variables such as age of onset, illness duration, medication exposure, or disease severity across disorders was beyond our scope. Furthermore, this study focused on adult neuroimaging data, and the nature and magnitude of case-control differences may vary as a function of age^12^. In future studies, harmonized multisite clinical protocols across different disorders, such as those informed by the Hierarchical Taxonomy of Psychopathology^85^, collected across the lifespan, could be used to investigate these effects in more detail.

Our cases were diagnosed according to DSM criteria^86^. Given the widespread application of DSM in clinical and research contexts, we deemed it important to understand heterogeneity with respect to these constructs and to gain insight into the neural correlates of phenotypic similarities and differences between cases assigned specific diagnoses. Nonetheless, focusing on specific syndromes rather than traditional diagnoses, as in the lesion network mapping literature^32^, may yield a more precise mapping between regional deviations, their network context, and behavior. Establishing sufficiently large databases for conducting such syndrome-focused analyses will be a key challenge for future work.

One factor complicating comparisons between regional, circuit, and network-level analyses is that the pipelines used for mapping deviation overlap rely on different thresholds. For instance, regional deviations are defined at z>|2.6|, whereas circuit-level results additionally rely on the threshold used to define the FC of deviant regions and the vertex-to-parcel mapping threshold (see Methods). In our analyses, we used a standard whole-brain corrected threshold for mapping the FC of deviant brain regions, so our results in this context align with typical expectations of the field. We also demonstrated that our findings were consistent across reasonable vertex-to-parcel mapping thresholds of 50% and 75%, suggesting a relative insensitivity to this parameter (see Figures 3, S11- 15). We also obtained similar results for the regional analysis when using an approach that integrates across a range of thresholds for defining extreme deviations that does not rely on a single fixed threshold value (See Methods, Figures S8-10).

In conclusion, our multiscale analysis of neural heterogeneity across six psychiatric disorders confirms that extreme regional heterogeneity of GMV deviations is a general characteristic of mental illness. Further, we showed that these deviations are often coupled to common functional circuits and networks, offering a putative neural substrate for phenotypic similarities between individuals assigned the same diagnosis. Similarly, the common involvement of prefrontal and parietal circuits, and the salience/ventral attention network, across disorders may be a marker of transdiagnostic psychological distress, with the variable involvement of other systems explaining phenotypic differences between disorders. As has been extensively documented in neurological syndromes^32^, our findings underscore the need to consider the network context of disorder-related pathophysiology^29^ and indicate that the clinical expression of disease is not driven solely by sites of primary pathology, but also by the effect of this pathology on remote, connected systems.

## MATERIALS AND METHODS

The study was approved by the local ethics committee of each dataset, and written informed consent was obtained from each participant. This study was approved by the Monash University Research Ethics Committee (Project ID: 23534).

### Participants

This study included 3746 individuals (1865 healthy controls, HC; 1833 cases across six different diagnostic categories) from 14 separate, independently acquired datasets and 25 scan sites. Full details on study design and clinical characteristics have been described previously for each dataset (see Table S1 for basic characteristics and relevant references). Each study was approved by the relevant ethics committee and written informed consent was obtained from each participant.

The final sample that we examined was taken from a larger pool of individuals recruited across the 14 datasets. In addition to the specific quality control procedures used within each study (references in Table 1), we performed a series of additional quality control checks and exclusions for our analysis. Specifically, we excluded participants if they were below 18 years or above 64 years of age (*N*=346); did not have the necessary clinical data (clinical diagnosis or, for healthy controls, absence of *any* clinical diagnosis) or demographic information (age, sex and scanner site, *N*=53); if their T1-weighted structural magnetic resonance imaging (MRI) scan did not survive our stringent manual and automated quality control procedure, as explained below (*N*=269); or if the data came from a site with less than 10 individuals in the same group and sex (described in the *Normative model* section, below; *N*=217). Our final sample available for analysis thus comprised 1465 HCs and 1294 cases. Demographic and other details of this cohort are presented in Table S1.

**Table 1.**
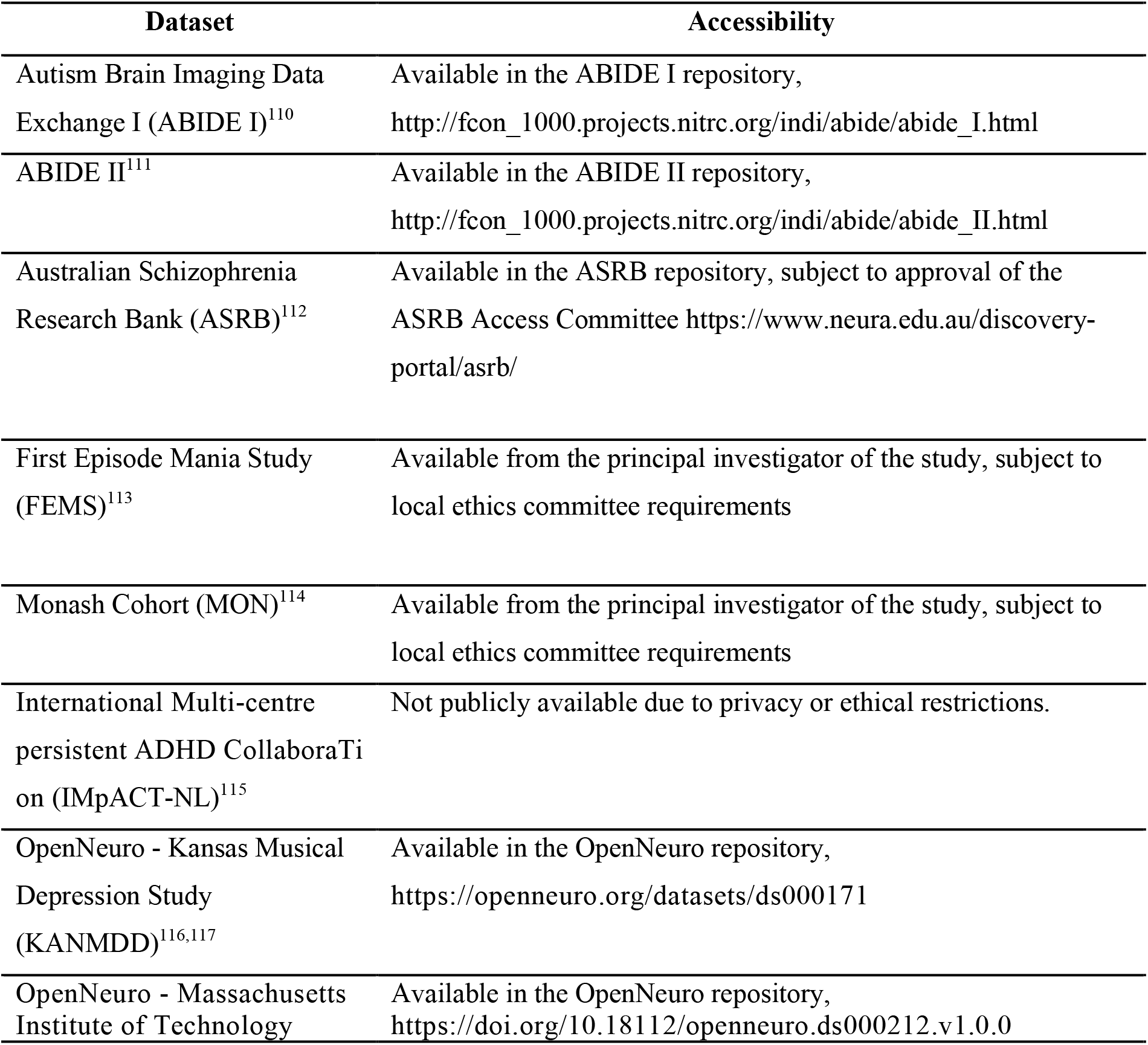
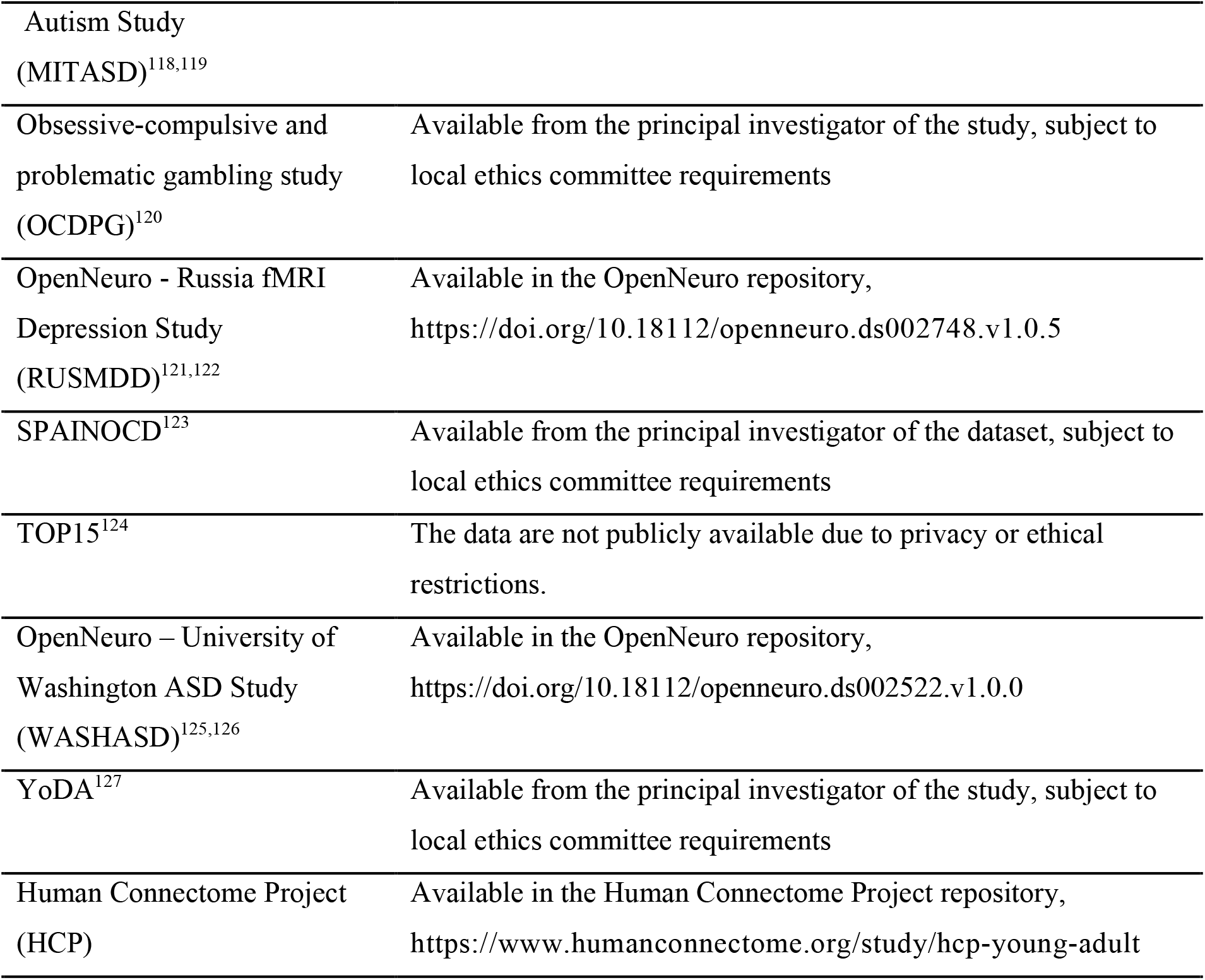
Summary of data availability for each dataset used in this study

### Mapping neural heterogeneity at the regional level

Our analysis aimed to characterize neural heterogeneity at the level of individual brain regions, neural circuits, and extended brain networks (Figure 1). To map heterogeneity at the regional level, we obtained person-specific deviation maps, representing the extent to which the grey matter volume (GMV) of a given person within each brain region deviates from normative predictions. This analysis involved quantifying regional GMV for each individual and evaluating the observed measures with respect to an underlying normative model, as outlined in the following.

#### Anatomical Data

We estimated regional GMV using voxel-based morphometry^87^ (VBM) of T1-weighted anatomical MRI scans^52,87^. We focused on GMV because it is one of the most frequently studied neural phenotypes in psychiatry. VBM is also arguably the most widely used tool for measuring GMV in this context. Image acquisition parameters for each dataset are provided in Table S1. The following outlines our quality assurance procedures and data processing pipeline, which was applied to all raw T1-weighted images obtained for each dataset.

#### Quality Control

All T1-weighted images were visually inspected and evaluated for the presence of artifacts^88,89^, resulting in the exclusion of 53 images with gross artifacts or abnormalities. Next, we used the Computational Anatomy Toolbox^52^ (CAT12 r113, http://dbm.neuro.uni-jena.de/cat/) to generate a weighted overall image quality rating (IQR) for every scan. This metric combines ratings of basic image properties, including the level of noise and geometric distortions, into a single score that quantifies the overall image quality of a participant’s T1-weighted scan (see here for more information www.neuro.uni-jena.de/cat/index.html#QA). On this metric, lower scores denote higher image quality. As per previous work^21^, we excluded 153 images with an IQR > 2.8. An additional 63 images were excluded due to a failure of our processing pipeline. Collectively, this quality assurance process resulted in the exclusion of a total of 269 scans.

#### MRI preprocessing

Regional GMV was estimated using the CAT12 VBM pipeline, which is included as an extension of Statistical Parametric Mapping software (SPM12, http://www.fil.ion.ucl.ac.uk/spm/software/spm12) in MATLAB v9.8^90^. Briefly, the T1-weighted images were first corrected for intensity nonuniformities, and segmented into GM, white matter (WM), and cerebrospinal fluid (CSF) tissue probability maps. Then, using the high- dimensional Diffeomorphic Anatomical Registration Exponentiated Lie Algebra^53^ (DARTEL), the segmented scans were normalized into standard Montreal Neurological Institute (MNI) space. Lastly, the images were bias-field corrected and modulated by the linear and nonlinear components of the Jacobian determinant obtained from the DARTEL deformation fields to obtain voxel-wise estimates of GMV. To constrain our analyses to GM voxels, we generated a mean image from all the normalized GM maps and retained voxels with a tissue probability ≥0.2.

#### Brain parcellation

We employed hierarchical Bayesian regression (HBR) to estimate normative models of regional GMV (see *Normative Model* section, below). To limit computational burden, we parcellated the brain into 1032 cortical and subcortical regions by combining well-validated parcellations of the cortex^54^ comprising, 1000 regions, and of the subcortex^55^, comprising 32 regions. The cortical parcels have been mapped to well-described, canonical functional networks of the brain^58^, facilitating our analysis of network-level overlap in deviations. Voxels that overlapped between atlases were assigned to the corresponding subcortical region. Regional GMV estimates were obtained using freely available code (http://www0.cs.ucl.ac.uk/staff/gridgway/vbm/get_totals.m).

Note that the choice of parcellation will necessarily affect the magnitude of overlap that can be observed in any given brain area, such that coarser parcellations will be associated with higher levels of overlap. Comparison to the control data thus provides a critical normative benchmark against which to evaluate the levels of overlap observed in each clinical group. We chose our 1032-region parcellation to offer as much spatial resolution as possible while still ensuring that our analyses were computationally feasible. The same parcellation was used in regional, circuit, and network-level analyses, facilitating direct comparison across scales.

#### Normative Modelling

We used normative modeling to obtain person-specific GMV deviation maps in relation to an underlying model of normative expectations for regional GMV variations (see ^27^ for details; code available at https://github.com/amarquand/PCNtoolkit, version=0.16). Normative models estimate the mean and variance (referred to as a normative range) of a response variable (e.g., GMV) from a set of clinically relevant covariates (e.g., age and sex) across a large healthy sample, referred to as a *training set*. These estimates are then used to quantify the deviations of samples in the *test subset*, which typically consists of cases, from the normative range.

When using multi-site data, scanner- and site-related variability introduce artefactual variance that confounds the result of any subsequent analyses^91^. These confounding effects present as site-correlated biases that cannot be explained by biological heterogeneity between samples. To account for these site-related effects, we built our normative model using hierarchical Bayesian regression (HBR), which successfully accommodates signal and noise variance in multisite data by estimating different but connected mean and variance components through shared prior distributions across sites^56^. Prior to modelling, regional GMV estimates were first subjected to a Box-Cox transformation to ensure normality^92^. The optimal lambda parameter for minimizing skewness was estimated for each brain region independently using maximum likelihood.

For the model to accurately parse variance attributable to age, sex, and site, a sufficient number of observations is required for any given combination of these variables. We therefore excluded data cells containing less than 10 HCs for a particular sex at a given site. If this entire exclusion procedure resulted in less than 10 cases in total from any given site, the entire site was excluded, resulting in the exclusion of 217 scans.

The training data for the normative model (HC_train_) was created by randomly selecting 90% of HC individuals from each site, provided that the site included data for ≥30 HCs. For sites with smaller HC samples, all HC data were included in the training set. The test data (HC_test_), which was completely independent of the normative model, comprised the 10% of HCs from sites with ≥30 HCs and all case scans. The HC_test_ data, comprising 269 individuals (140 male), offered a normative benchmark for assessing case-specific model deviations, as outlined below.

Following stratification of our sample into training and test subsets, for each brain region, we fitted separate HBRs^56^, modeling site and sex effects with random slopes, intercepts, and noise, to each of our 1032 parcellated brain regions to model GM volume as a function of age, sex, and site in the training data, yielding estimates of normative regional GMV variance and predictive uncertainty. Importantly, based on a partial-pooling approach, shared prior distributions were imposed over site- specific and sex-specific model parameters. These shared priors assume that while the model parameters for each site and sex are different, they are drawn from a common distribution. These shared priors thus regularize the model parameters and prevent the model from overfitting small batches for a given sex or site.

To quantify deviations from the normative model predictions for each subject in the test data, we generated deviation *z-*maps for each individual. Specifically, for each subject, *i*, at each brain region, *j*, we combined the predicted GM volume, 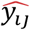, true GM volume, *y*_*ij*_ the predictive uncertainty, *σ*_*ij*_ and the normative variance, *σ*_*nj*_, to calculate a z-score, *z*_*ij*_ as

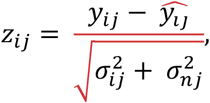

which quantifies the extent to which an individual’s regional GMV estimate deviates from the model prediction, given the uncertainty of the model.

For any given brain region, we are interested in participant values that deviate to a large extent from the normative expectations set by the model, under the assumption that these deviations represent pathophysiologically relevant features of the disease^27^. We used two complementary approaches to characterize positive and negative deviations from the normative model at each region and for each individual. First, we thresholded the deviation maps at |z| > 2.6 (i.e., *p* < 0.005) to identify regions showing extreme deviations from model predictions. We used this approach to be consistent with prior work^21,93^ and because alternative methods, such as the False Discovery Rate^94^ (FDR), rely on adaptive thresholds that can vary between individuals. Applying a fixed *z*-threshold allows for an absolute definition of extreme deviations. Second, to ensure that our findings were not driven by this specific choice of threshold, we adapted a threshold-weighted approach^57^, as detailed below.

#### Evaluation of model performance

To assess model generalizability, we used 5-fold cross-validation (CV) applied to the HC_train_ cohort (*n* = 1196 HC individuals, 55% male). Specifically, we split this group into 5 folds. Within each fold, we estimated an HBR model on 80% of participants, using age, sex, and site as covariates, to predict regional GMV in the remaining 20% of people. This procedure was repeated 5 times so that regional GMV values for all participants in the HC_train_ group were predicted once. As above, we estimated deviation z-maps for every individual and identified extreme deviations.

For each fold in the CV, we assessed model fit for each brain region by evaluating three performance metrics: (i) explained variance (EXPV); (ii) the mean standardized log-loss (MSLL); and (iii) the standardized mean-squared error (SMSE)^56^, as shown in Figure S3. We also evaluated the model’s efficacy in partitioning site-related variance in the data using linear support vector machines (SVM). Specifically, we used a series of one-versus-all linear support vector machines (LSVM with default slack parameter = 1) trained separately on the z-maps from the HC_train_ and the HC_test_ subsets to classify scan sites. For each site, we ran a 2-fold SVM classifier to obtain the mean balanced accuracy score for the given site. Here, a balanced accuracy at chance-level (50%) indicated that the resulting deviations were not contaminated by residual site effects, as confirmed in Table S2. To assess whether individual variations in scan quality affected the deviation z-maps, we calculated the Pearson’s correlation between the total number of extrema and CAT12’s IQR rating in each group.

#### Characterizing the regional heterogeneity of extreme deviations

We characterized the heterogeneity of case-specific thresholded deviation maps for each disorder using a non-parametric approach. Specifically, for each clinical group and the HC_test_ cohort, we computed the proportion of individuals in each group showing an extreme deviation within each region, resulting in group-specific overlap maps estimated separately for positive and negative extrema (Figure 2f). We used the proportion of individuals rather than raw counts to account for sample size differences between groups. Next, we subtracted the HC_test_ overlap map from each disorder’s overlap map, resulting in an overlap difference map for each disorder (Figure 2g). We then permuted group labels (i.e.,HC, case) and repeated the procedure 10,000 times to derive an empirical distribution of overlap difference maps under the null hypothesis of random group assignment (Figure S6a-c). For each brain region, we obtained *p*-values as the proportion of null values that exceeded the observed difference (Figure S6d). The tails of the null distribution (i.e., values associated with *p*< 0.10) were approximated using a generalized Pareto distribution^95^, as implemented in the Permutation Analysis of Linear Models software package^96^ (PALM alpha116), to allow inference at arbitrarily high levels of precision. Statistically significant effects were identified using an FDR-corrected^94^ threshold of *p*_*FDR*_<.05, two-tailed.

#### Threshold-weighted deviation mapping

The procedure to characterize the regional heterogeneity of extreme deviations above used an arbitrary threshold (Z< |2.6|) on each individual z-map. The practical benefit of this approach is that it allows calculation of intuitive metrics, such as the proportion of cases within a group showing a supra-threshold deviation within a given brain region. We complemented this analysis with an alternative approach, which does not yield similarly intuitive metrics but which integrates results across a range of thresholds^57^, thus allowing us to determine the degree to which our findings depend on a specific threshold choice. First, for each diagnostic group, we thresholded z-maps across the threshold range 1.64 < *Z* < 3.10, in 100 equal log-space increments, separately for positive and negative extrema. Second, we obtained a region- and threshold-specific percentage overlap map quantifying the proportion of individuals within each group showing an extreme deviation for any given region and threshold. Third, we applied a weighted function to penalize less conservative thresholds. Fourth, we obtained a final threshold-weighted overlap map by taking the area under the curve of the cumulative histogram of the proportion of participants at each threshold for each region across participants. We used the following linear weighted function proposed by Seghier and Price^57^, which ensures that overlap map values range between 0 and 1, with 1 indicating an effect in a region is present in each subject at each threshold:

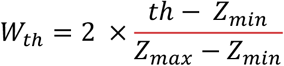

where *Z*_*max*_ and *Z*_*min*_ respectively correspond to the maximum and minimum z-value thresholds, and *th* is the given statistical threshold. We then calculated the difference between the threshold-weighted percentage overlaps between cases and the held-out HC cohort, resulting in an overlap difference map for each disorder. We performed inference on these differences using the same group-based permutation procedure described above with statistically significance defined using a threshold of *p*_*FDR*_<.05, two tailed.

### Mapping neural heterogeneity at the circuit level

Having investigated neural heterogeneity at the level of individual brain regions, we next evaluated whether regions showing extreme deviations in individual cases are functionally coupled to common areas, which we term circuit-level heterogeneity (Figure 1 middle). We define the functional circuitry of a brain region showing an extreme deviation as the set of brain areas that show significant FC with that region in an independent cohort of healthy individuals. To map this circuitry, we adapted elements of the lesion network mapping methodology used extensively in neurological disorders^31,32^ for use with normative modelling, as outlined in the following.

#### Functional magnetic resonance imaging (fMRI) data and processing

The functional circuitry of deviant regions was mapped using an independent cohort of 150 resting-state functional MRI (rs-fMRI) scans (71 males, age 21- 35 years) from the S900 release of the Human Connectome Project^97^ (HCP). These individuals corresponded to the 150 people with the lowest total head motion, as estimated using framewise displacement^98^, from the broader set of 282 unrelated participants with the same fMRI reconstruction in the S900 release. We focused on this sub- sample to minimize the effects of head motion in our FC maps and to minimize computational burden due to the large number of analyses required. Mapping deviation-related functional circuity in an independent cohort is essential because we seek to understand the circuit context of deviant regions under normative conditions. Our approach aligns precisely with the logic used in lesion network mapping of neurological patients^31,32^.

The rs-fMRI data for each subject consists of four runs (rfMRI_REST1_LR, rfMRI_REST1_RL, rfMRI_REST2_LR, rfMRI_REST2_RL) acquired at two different sessions (REST1 and REST2) using two different directions of phase coding (LR: left to right, and RL: right to left). Here, we mapped deviation-related FC using the rfMRI_REST1_LR data. All rs-fMRI data were obtained using a 32-channel Siemens 3T connectome-Skyra scanner. The imaging parameters for rs-fMRI were as follows: repetition time (TR) = 720 ms, echo time (TE) = 33.1 ms, flip angle (FA) = 52°, field of view (FOV) = 208 × 180 mm^2^, matrix = 104 × 90, slice number = 72, slice thickness = 2 mm, voxel size = 2 × 2 × 2 mm^3^, multiband factor = 8, and 1200 volumes. Participants were required to stay awake, relaxed, and to keep their eyes open and fixed on a bright cross-hair projected on a dark background. A detailed description of the HCP data is available in previous work^97^.

The rs-fMRI data used underwent the HCP’s minimal pre-processing pipeline^99^, which includes gradient-nonlinearity-induced distortion, motion correction to the single-band reference image using FLIRT, EPI image distortion correction using TOPUP^100^, registration into standard space using a customized boundary-based-registration (BBR) algorithm, and single step spline interpolation using all transforms, intensity normalization and bias field removal to resample the original EPI into MNI space. Minimal high pass filtering was applied with a cutoff of 2000ms. Artifacts were then removed using ICA-FIX^101^. This involves employing an automatic classifier, specifically trained for HCP data, to identify ICA components due to measurement noise, additional motion or physiological artifacts like cardiac pulsation and respiration. Next, the volume timeseries were mapped into the standard CIFTI grayordinate space^97^ and smoothed to 2mm FWHM (where the smoothing was on the surface for the cortex and in volume space for subcortex). This results in a standard set of grayordinates in every subject, with surface vertex data and subcortical volume voxel data. The mean GM signal was then removed each grayordinate’s time series to remove residual widespread signal deflections that are not removed by ICA-FIX^102–104^

#### Mapping the functional circuitry of extreme deviations

We used seed-based FC analysis to map the functional circuitry of each region showing an extreme deviation in any individual within the clinical and HC_test_ groups. Specifically, each deviant region was used as a seed, from which the average time course was extracted for each individual in the HCP_150_ sample (Figure 4b). These seed time courses were then correlated with all other brain grayordinates and the resulting correlation maps were subjected to Fisher’s *r*-to-*z* transformation^105^. The transformed correlation maps for each HCP_150_ subject were then aggregated using a one-way *t*- test at each grayordinate, as implemented in PALM. Thresholded maps representing the functional circuitry of the seed were obtained using permutation testing (500 permutations) with a generalized Pareto approximation of the tail of the null distribution^95^ and threshold-free cluster enhancement^106^ (TFCE), run separately for the cortical and sub-cortical areas, with a threshold of p_FWE_<.025, family-wise error corrected (i.e., .05/2 to account for cortical and subcortical analyses). This procedure was repeated for each region in which at least one extreme deviation across individuals was observed (Figure 3b).

To quantify circuit-level overlap, we developed an approach that parallels traditional lesion network mapping methodology but which allows statistical inference on observed case-control differences in overlap scores at each region. Specifically, we thresholded each deviant-related FC map by applying TFCE to the grayordinate maps, as described above. We then classified an area in our 1032-region parcellation as showing significant FC with the seed if more than 50% of its grayordinates survived the TFCE threshold. We repeated our analysis using a mapping threshold of 75% to ensure that our findings were not driven by this specific choice (see Figure S13-15). We mapped FC at the grayordinate level and thresholded the data in this way to leverage the superior statistical sensitivity of TFCE and to map the spatial architecture of the seed-related FC patterns more accurately. Our procedure resulted in a binary map representing the specific brain regions that comprise the functional circuitry of each seed (Figure 3c). Then, for each individual, we took the union of the binary FC maps across that person’s set of extrema, resulting in a single map identifying areas showing significant FC to any extreme deviation expressed by the person (Figure 3d). Separate union maps were obtained for positive and negative deviations. The overlap of these union maps was then taken for each clinical group and the HC_test_ sample, representing, for each region, the proportion of individuals in that group for which significant deviant-related FC was identified. We refer to this image as a circuit-level overlap map (Figure 3e).

Note that our approach focuses on FC between deviant and other areas as derived from a normative sample. It is possible that patterns of FC are altered in diagnosed individuals, and that the underlying network architecture differs from normative expectations. The net effect of such alterations would be to affect coupling between deviant and other areas, and thus our normative benchmark still offers an important reference point for understanding the distributed effects of regional deviations. Further investigation of how altered patterns of FC may moderate these relationships remains an important topic for future investigation.

#### Statistical inference on group differences in circuit-level overlap

We evaluated group differences in the circuit-level overlap maps of each clinical group relative to the HC_test_ group in two ways (see Figure S6 for a schematic overview). First, following the analysis of regional overlaps, we computed the difference in the case and control overlap maps (Δ *overlap map*, Figure 3e) and evaluated the statistical significance of these differences with respect to an empirical null distribution generated by permuting the group labels of the individual-specific FC union maps 10,000 times, with a generalized Pareto Tail approximation^95^. Statistically significant differences in deviation FC overlap were identifying using a threshold of *p*_*FDR*_ < 0.05, two-tailed (Figure S6a-d).

This group-based permutation test identifies regions where the overlap in deviant-related functional circuitry differs between cases and controls, but it does not preserve group differences in the total number of deviations. It therefore cannot distinguish whether any observed differences arise from a preferential aggregation of deviations within specific circuits in one group relative to the other, or whether the group differences are driven by variations in total deviation burden. Note that differences in deviation burden should not be thought of as a confound in this context, since they are an intrinsic feature of the disease that will have real phenotypic consequences. Nonetheless, it is important to determine which findings might be driven by differences in deviation burden compared to a preferential accumulation of pathology within a specific circuit for a given disorder.

To disentangle these possibilities, we considered a second, spatial null model that preserves group differences in deviation burden. Specifically, we generated an ensemble of null cortical deviation maps for each individual in the test data by spatially rotating their empirical, unthresholded deviation maps (Figure S6e) using Hungarian spherical spin tests^107,108^ (Figure S6f) and then thresholding these rotated maps in the same way as the observed data (i.e., Z> |2.6|; Figure S6g). The Hungarian method was used as it preserves the original values and spatial autocorrelation of the original map, which ensures that the surrogate data yields the same number of extrema as originally observed. This procedure could only be applied to the cortex, which is topologically equivalent to a sphere. The choice of an appropriate null model for subcortical areas is more complicated. While model-based procedures (e.g.,^109^) can generate surrogates with comparable spatial autocorrelation, they require parameter tuning, can show variable fits across individuals, and do not preserve the exact values of the original data, making thresholding challenging. We therefore randomly shuffled deviation values across subcortical areas before thresholding them. Note that this null model is more lenient than a spatially constrained test since the model preserves fewer features of the data. As we found no significant differences using this test, our results will not change under a more stringent, spatially constrained null model.

We then obtained individual-specific surrogate FC union maps using the same procedure described above (Figure S6h), and calculated surrogate within-group overlap maps (Figure S6i) and between-group overlap difference maps (Figure S6j). This procedure was repeated 10,000 times to generate a null distribution of overlap difference maps for each disorder, again using the generalized Pareto tail approximation (Figure S6k). Statistically significant differences were identified using a threshold of *p*_*FDR*_ < 0.05, two-tailed.

To summarize, group differences in circuit-level overlap were assessed with respect to two null models, one based on permutation of group labels and one based on spatial permutation of individual deviation maps. The group-based permutation test identifies regions showing differences in circuit-level overlap regardless of deviation burden. The spatial permutation test can be used to assess the degree to which such differences are driven by variations in deviation burden. More specifically, if a given brain region shows a significant difference for both tests, we have evidence to indicate that extreme deviations within the disorder preferentially aggregate within the functional circuitry of that region. If, on the other hand, a region shows a difference with the group-based permutation test but not the spatial permutation test, then the observed group differences are consistent with expectations from randomly selecting of the same number of seeds in each group, indicating that the effects observed under group-based permutation can be attributed to the fact that one group has a higher deviation burden than the other. Note that this scenario still yields important information about the extent of circuit-level heterogeneity in the disorder, since we generally expect cases to have a higher deviation burden and any resulting circuit-level overlap will still have phenotypic consequences regardless of whether such overlaps are explained by deviation burden or not. The spatial permutation test simply offers additional insights into the potential mechanisms that may drive group differences in circuit-level overlap.

### Mapping neural heterogeneity at the network level

The above procedure offers a means for understanding heterogeneity at the level of neural circuits that show strong FC with a deviant region. However, it is still possible for some pairs of brain regions to be affiliated with the same extended functional network despite being weakly coupled themselves (Figure 1 right). We therefore sought to characterize neural heterogeneity at the level of the broader functional networks within which a given deviant region may be embedded.

#### Assigning regions to networks

We assigned each cortical region to one of 7 canonical functional cortical networks using a well-validated network parcellation^54,58^ and assigned each subcortical region to either the medial temporal lobe (amygdala and hippocampus), thalamus, or basal ganglia (nucleus accumbens, globus pallidus, putamen, caudate nucleus), as done previously^55^, resulting in a total of 10 distinct functional networks (Figure 4a-c). The subcortical regions were assigned to coarse anatomical areas that were not aligned with the finely mapped cortical function networks because the correspondence between these subcortical nuclei, as parcellated here, and canonical cortical functional networks has not been extensively investigated. An alternative network assignment may influence network-level subcortical overlap values.

#### Characterizing network-level heterogeneity

To characterize network-level heterogeneity, we examined the degree to which GMV deviations aggregated within each network. To this end, for each network, we estimated the proportion of individuals within each diagnostic group that showed at least one extreme deviation in a region assigned to that network, separately for positive and negative deviations (Figure 4d). We then computed clinical-control differences in overlap proportions for each network (Figure 4e).

#### Statistical inference on group differences in network-level overlap

As in the circuit-level analysis, these observed group differences in network-level overlap were evaluated with respect to two complementary null models. The first involved a group-based randomization test in which a null distribution was generated by permuting group labels 10,000 times (Figure S6a-d). The second null model used the 10,000 spatially rotated maps generated in the circuit-level analysis to obtain surrogate estimates of group differences in network-level overlap while preserving variation in deviation burden (Figure S6e-g,l-o). In both cases, the tail of the null distribution was approximated using a generalized Pareto distribution and we used a significance threshold of *p*_*FDR*_ < 0.05, two-tailed. As in the circuit-level analysis, these two null models allowed us to distinguish preferential network involvement from the effects of total deviation burden. Note that comparisons in overlap values *between* networks are complicated by differences in network size. Our analysis therefore focuses primarily on comparisons between cases and controls *within* networks since the control data provide a critical normative benchmark against which cases can be compared.

## Data Availability

Raw data for the ABIDE studies are available online at http://fcon_1000.projects.nitrc.org/indi/abide/. Raw data for the ASRB study is available in the ASRB repository, subject to the approval of the ASRB Access Committee https://www.neura.edu.au/discovery-portal/asrb/. Raw data from the FEMS, MON, OCDPG, SPAINOCD, and YoDA studies are available from the principal investigator of the dataset, subject to local ethics committee requirements. Raw data from the KANMDD, MITASD, RUSMDD, WASHASD studies are available from the OpenNeuro repository. Raw data for the HCP study is available online at https://www.humanconnectome.org/study/hcp-young-adult. Raw data for IMpACT-NL and TOP15 are not publicly available due to privacy or ethical restrictions.

## Data availability

## Code availability

All code used to produce the figures in this study are freely available online and can be found at https://github.com/ashlea-segal/multiscale-heterogeneity-brain-abnormalities.

## Acknowledgements

This work was supported by the MASSIVE HCP facility (http://www.massive.org.au). This study was supported by the Australian Schizophrenia Research Bank (ASRB), which is supported by the National Health and Medical Research Council of Australia, the Pratt Foundation, Ramsay Health Care, the Viertel Charitable Foundation and the Schizophrenia Research Institute. This research was supported by grants from the European Research Council (ERC, grant ‘MENTALPRECISION’ 10100118 and ‘BRAINMINT’ 802998), the Wellcome Trust under an Innovator award (‘BRAINCHART,’ 215698/Z/19/Z) and a Strategic Award (098369/Z/12/Z), the Dutch Organisation for Scientific Research (VIDI grant 016.156.415) the Research Council of Norway (223273, 249795, 298646, 300768, and 276082), the South-Eastern Norway Regional Health Authority (2014097, 2015073, 2016083, and 2019101), the Carlos III Heath Institute (PI16/00889 and PI19/01171), the Generalitat de Catalunya (PERIS grant SLT006/17/00249). The work was also supported by grant U54 EB020403 to the ENIGMA Consortium from the BD2K Initiative, a cross-NIH partnership, and by the European College of Neuropsychopharmacology (ECNP) Network “ADHD Across the Lifespan”, and by the Australian National Health and Medical Research Council of Australia (NHMRC) Project Grants 1024570 (principal investigator, CGD) and 1064643 (principal investigator, BJH). LP was supported by the National Institute Of Mental Health of the National Institutes of Health under Award Number K99MH127296 and a 2020 NARSAD Young Investigator Grant from the Brain & Behavior Research Foundation. The content is solely the responsibility of the authors and does not necessarily represent the official views of the National Institutes of Health. TW gratefully acknowledges the European Union’s Horizon 2020 research and innovation programme under the Marie Sklodowska-Curie grant agreement No 895011. BF is supported by a personal Veni grant from the Netherlands Organization for Scientific Research (NWO, grant number 016–130-669). MH is supported by a personal Veni grant from the Netherlands Organization for Scientific Research (NWO, grant number 91619115). AZ was supported by a research fellowship from the NHMRC (APP1118153). CGD and BJH were supported by NHMRC Career Development Fellowships (1141738 and 1124472, respectively). JT was supported by the Turner Impact Fellowship from the Turner Institute for Brain and Mental Health. MY role on this paper was funded through a National Health and Medical Research Council Fellowship (NHMRC; #APP1117188). MY also receives funding from other NHMRC schemes, Monash University, and Australian Government funding bodies such as the Australian Research Council (ARC), Australian Defence Science and Technology (DST), and the Department of Industry, Innovation and Science (DIIS). MB was supported by a NHMRC Senior Principal Research Fellowship (1156072). SC was supported by a NHMRC Senior Research Fellowship (<u>APP1136344</u>). MAB was supported by National Health and Medical Research Council (Senior Research Fellow - Level B). AF was support by the Sylvia and Charles Viertel Foundation, National Health and Medical Research Council (IDs: 1197431, 1146292, and 1050504) and Australian Research Council (ID: DP200103509). We would like to thank Seghier, M. L for sharing code to generate threshold-weighted overlap maps. We would like to thank all the sites and investigators who have worked to share their data through data sharing repositories including ABIDE and OpenNeuro. Additional data were provided [in part] by the Human Connectome Project, WU-Minn Consortium (Principal Investigators: David Van Essen and Kamil Ugurbil; 1U54MH091657) funded by the 16 NIH Institutes and Centers that support the NIH Blueprint for Neuroscience Research; and by the McDonnell Center for Systems Neuroscience at Washington University.

## Ethics declarations

KA is a scientific advisor to and shareholder in BrainKey Inc., a medical image analysis software company. BF has received educational speaking fees from Medice GmbH. CFB is director and shareholder of SBGNeuro Ltd. OAA is a consultant to HealthLytix and received speaker’s honorarium from Lundbeck and Sunovion. MY has received philanthropic donations from the David Winston Turner Endowment Fund, Wilson Foundation, as well as payments in relation to court- , expert witness-, and/or expert review-reports. Finally, he has received funding to conduct sponsored Investigator-Initiated trials (including Incannex Healthcare Ltd). These funding sources had no role in the design, management, data analysis, presentation, or interpretation and write-up of the data. MY also sits on the Advisory Boards of Centre of The Urban Mental Health, University of Amsterdam; Enosis Therapeutics; and Monash Biomedical Imaging Centre. MB has received Grant/Research Support from the NIH, Cooperative Research Centre, Simons Autism Foundation, Cancer Council of Victoria, Stanley Medical Research Foundation, Medical Benefits Fund, National Health and Medical Research Council, Medical Research Futures Fund, Beyond Blue, Rotary Health, A2 milk company, Meat and Livestock Board, Woolworths, Avant and the Harry Windsor Foundation, has been a speaker for Abbot, Astra Zeneca, Janssen and Janssen, Lundbeck and Merck and served as a consultant to Allergan, Astra Zeneca, Bioadvantex, Bionomics, Collaborative Medicinal Development, Eisai, Janssen and Janssen, Lundbeck Merck, Pfizer and Servier – all unrelated to this work. MB has received grant/research support from National Health and Medical Research Council, Wellcome Trust, Medical Research Future Fund, Victorian Medical Research Acceleration Fund, Centre for Research Excellence CRE, Victorian Government Department of Jobs, Precincts and Regions and Victorian COVID-19 Research Fund. He received honoraria from Springer, Oxford University Press, Cambridge University Press, Allen and Unwin, Lundbeck, Controversias Barcelona, Servier, Medisquire, HealthEd, ANZJP, EPA, Janssen, Medplan, Milken Institute, RANZCP, Abbott India, ASCP, Headspace and Sandoz. The other authors report no conflicts of interest.

