## Supplementary Material for "Regional, circuit, and network heterogeneity of brain abnormalities in psychiatric disorders"

**SUPPLEMENTARY MATERIALS**

*Table S*1. Demographic and scanner details of the patient and control groups

| Dataset | DX Groups | Sites Included | Scanner | Voxel Size ($mm^3$) | Field Strength | $HC_{train}$ | | $HC_{test}$ | | Patients | |
|---|---|---|---|---|---|---|---|---|---|---|---|
| | | | | | | No subjs (% Male) | Age (year), median (SD) [range] | No subjs (% Male) | Age (year), median (SD) [range] | No subjs (% Male) | Age (year), median (SD) [range] |
| Autism Brain Imaging Data Exchange I (ABIDE I)[1] | Autism Spectrum Disorder (ASD) | CALTECH[a] | Siemens Magnetom Trim Trio | 1 | 3T | 14 (100) | 26 (11) [19-56] | - | - | 14 (100) | 23 (10) [18-55] |
| | | CMU[b] | Siemens Magnetom Verio | 1 | 3T | 10 (100) | 26 (5) [21-40] | - | - | 11 (100) | 24 (5) [20-39] |
| | | Leuven_1[c] | Philips Intera | 0.98 x 0.98 x 1.2 | 3T | 14 (100) | 22 (2) [18-29] | - | - | 14 (100) | 20 (3) [18-32] |
| | | MaxMun[d] | Siemens Magnetom Verio | 1 | 3T | 19 (100) | 27 (7) [21-48] | 4 (100) | 26 (3) [23-32] | 13 (100) | 31 (11) [18-58] |
| | | NYU[e] | Siemens Magnetom Allegra | 1.3 x 1.3 | 3T | 20 (100) | 22 (3) [19-31] | 5 (100) | 25 (5) [18-32] | 15 (100) | 23 (5) [18-39] |
| | | PITT[f] | Siemens Magnetom Allegra | 1.1 x 1.1 x 1.1 | 3T | 11 (100) | 25 (5) [19-33] | - | - | 12 (100) | 23 (5) [18-35] |
| | | SBL[g] | Philips Intera | 1 | 3T | 14 (100) | 36 (5) [26-42] | - | - | 14 (100) | 31 (6) [22-49] |
| | | USM[h] | Siemens Magnetom Trim Trio | 1 x 1 x 1.2 | 3T | 24 (100) | 24 (6) [18-39] | 5 (100) | 25 (3) [19-27] | 40 (100) | 23 (7) [18-50] |
| ABIDE II[2] | ASD | BNI[i] | Philips Ingenia | 1.1 x 1.1 x 1.2 | 3T | 23 (100) | 43 (14) [18-64] | 6 (100) | 44 (14) [21-62] | 29 (100) | 41 (15) [18-62] |

| | | | | | | | | | | | |
|---|---|---|---|---|---|---|---|---|---|---|---|
| | | IU[j] | Siemens Magnetom Trim Trio | 0.7 x 0.7 x 0.7 | 3T | 14 (100) | 22 (5) [20-37] | - | - | 13 (100) | 20 (5) [18-37] |
| Australian Schizophrenia Research Bank (ASRB)[3] | Schizophrenia (SCZ) | BRIS | Siemens Avanto | 0.98 x 0.98 x 1 | 1.5T | 27 (48.15) | 41 (14) [18-63] | 6 (50) | 39 (8) [30-51] | 59 (71.19) | 37 (11) [20-64] |
| | | MELB | Siemens Avanto | 0.98 x 0.98 x 1 | 1.5T | 46 (50) | 35 (12) [19-61] | 11 (54.55) | 34 (12) [20-56] | 49 (63.27) | 35 (9) [20-54] |
| | | PERT | Siemens Avanto | 0.98 x 0.98 x 1 | 1.5T | 24 (45.83) | 33 (12) [18-59] | - | - | 16 (81.25) | 33 (7) [23-52] |
| | | SYDN | Siemens Avanto | 0.98 x 0.98 x 1 | 1.5T | 25 (48) | 41 (15) [18-62] | 6 (50) | 38 (8) [20-45] | 41 (63.41) | 39 (10) [20-64] |
| First Episode Mania Study (FEMS)[4] | Bipolar Disorder (BP) | | Siemens Magnetom Trim Trio | 1 | 3T | 24 (45.83) | 22 (1) [19-25] | 3 (0) | 21 (2) [19-24] | 38 (76.32) | 21 (2) [18-26] |
| Monash Cohort (MON)[5] | HC Only | | Siemens Magnetom Skyra | 1 | 3T | 315 (42.54) | 22 (5) [18-50] | 80 (42.5) | 23 (5) [18-41] | - | - |
| International Multi-centre persistent ADHD CollaboraTion | Attention Deficit Hyperactivity Disorder (ADHD) | | Siemens Magnetom Avanto | 1 | 1.5T | 116 (43.97) | 33 (11) [19-63] | 30 (43.33) | 28 (13) [20-61] | 153 (41.18) | 34 (10) [18-61] |

| | | | | | | | | | | | |
|---|---|---|---|---|---|---|---|---|---|---|---|
| (IMpACT-NL)[6] | | | | | | | | | | | |
| OpenNeuro - Kansas Musical Depression Study (KANMDD)[7,8] | Major Depressive Disorder (MDD) | | Siemens Magnetom Skyra | 1 x 1 x 1.2 | 3T | 11 (0) | 25 (10) [18-59] | - | - | 11 (0) | 26 (11) [18-52] |
| OpenNeuro - Massachusetts Institute of Technology. Autism Study (MITASD)[9,10] | ASD | | Siemens Magnetom Trim Trio | 1.33 x 1 x 1 | 3T | 14 (100) | 24 (9) [20-45] | 3 (100) | 25 (13) [20-50] | 11 (100) | 36 (7) [21-46] |
| Obsessive-compulsive and problematic gambling study (OCDPG)[11] | Obsessive-compulsive Disorder (OCD) | | Siemens Magnetom Skyra | 1 | 3T | 31 (48.39) | 31 (9) [19-54] | 7 (57.14) | 31 (6) [25-44] | 33 (48.48) | 29 (9) [19-53] |
| OpenNeuro - Russia fMRI | MDD | | Philips Ingenia | 1 | 3T | 12 (0) | 32 (8) [22-52] | 3 (0) | 26 (10) [23-47] | 37 (0) | 30 (9) [19-55] |

| | | | | | | | | | | | |
|---|---|---|---|---|---|---|---|---|---|---|---|
| Depression Study (RUSMDD)[12,13] | | | | | | | | | | | |
| SPAINOCD[14] | OCD | | GE Signa Excite | 1.17 x 1.17 x 1.2 | 1.5T | 110 (54.55) | 33 (9) [18-61] | 28 (53.57) | 33 (9) [19-60] | 134 (50.75) | 35 (9) [18-58] |
| TOP15[15] | SCZ, BP | | Siemens Magnetom Sonata | 1.33 x 0.94 x 1 | 1.5T | 203 (54.68) | 33 (9) [18-59] | 53 (54.72) | 33 (9) [18-53] | 218/190 (57.80/41.58) | 31 (9) [19-62] / 32 (11) [18-64] |
| OpenNeuro – University of Washington ASD Study (WASHASD)[16,17] | ASD | | Philips Achieva | 1 | 3T | 13 (100) | 21 (2) [18-26] | 3 (100) | 22 (2) [20-25] | 16 (100) | 22 (3) [18-30] |
| YoDA[18] | MDD | | GE Signa Excite | 0.94 x 0.94 x 1 | 3T | 62 (43.55) | 21 (2) [18-23] | 16 (43.75) | 20 (2) [18.25] | 113 (48.67) | 21 (2) [18.26] |

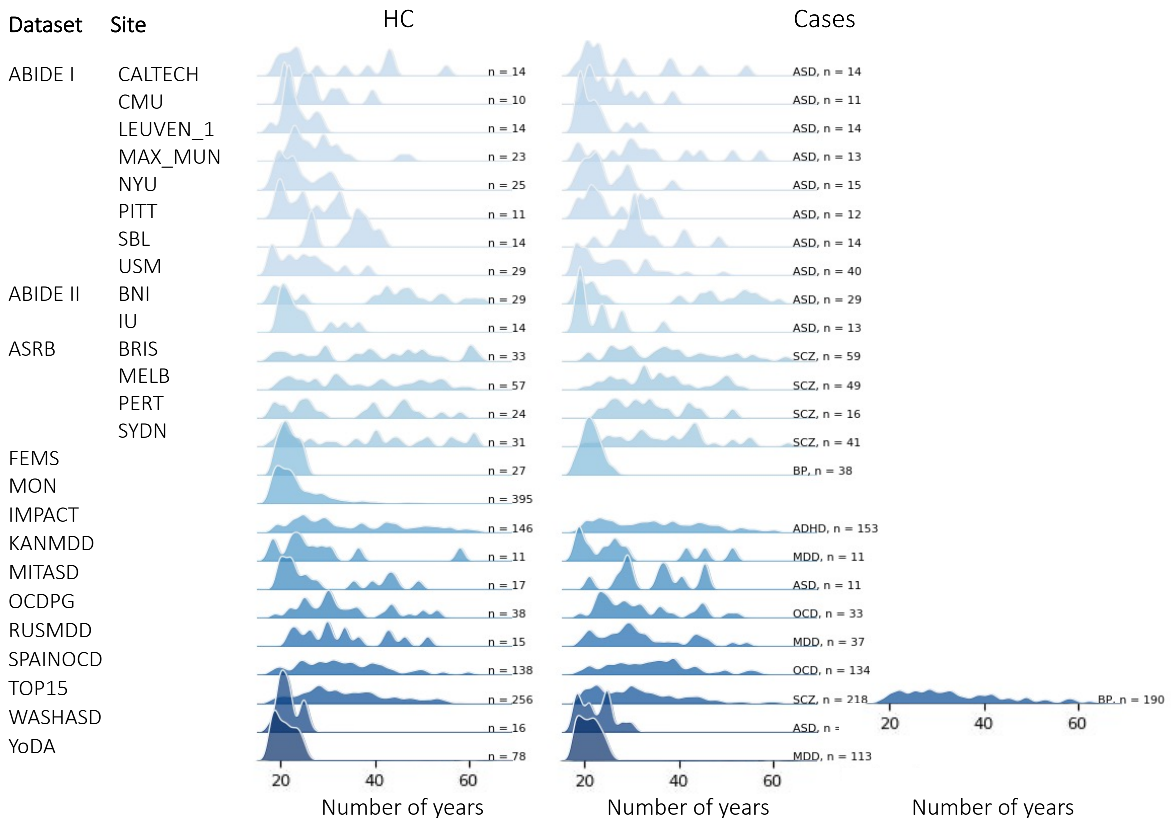


*Figure S1.* **Age distributions across scan sites for each diagnostic group**.

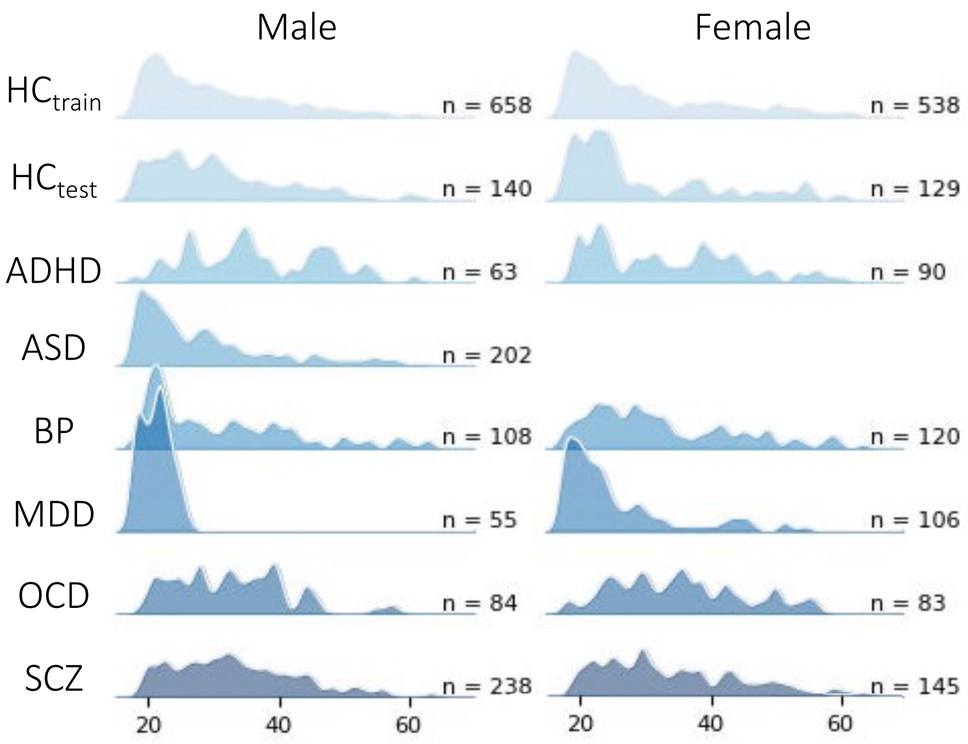


*Figure S2.* **Age distributions across diagnostic groups for each sex.**

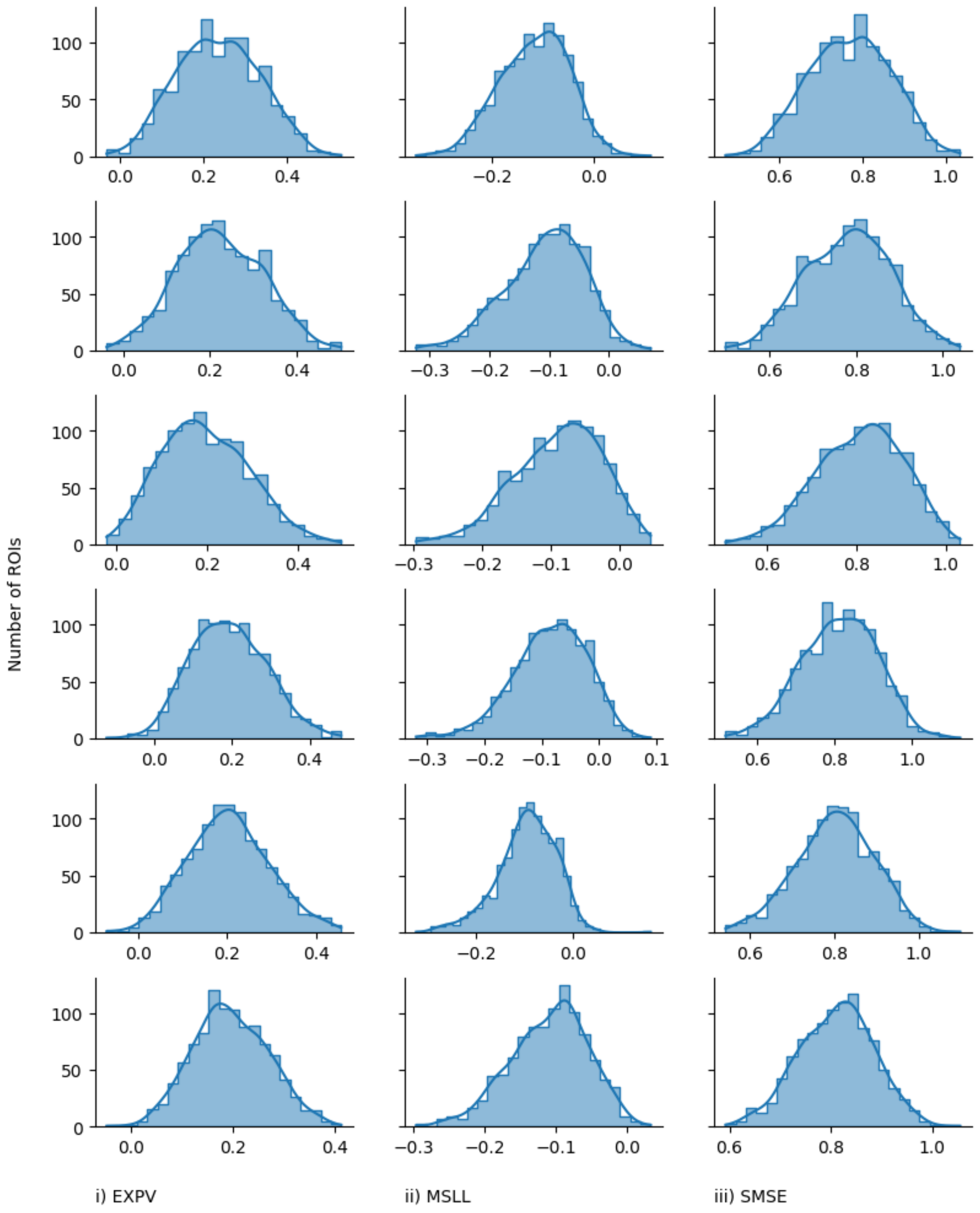


*Figure S3.* **Performance metrics for the normative models.** These metrics measure the accuracy with which our normative model estimated the relationship between GMV and age, sex, and site. The distributions of i) explained variance (EXPV; higher is better), ii) Mean standardized log-loss (MSLL; lower is better), and iii) Standardized mean squared error (SMSE; lower is better) across 1000 cortical and 32 sub-cortical regions in each HC CV sample ($HC_{train}$). The top five rows present data from each cross-validation fold. The bottom row presents data for the test cohort.

*Table S2.* Balanced classification accuracy scores (%) from the linear support vector machine within HC individuals from the CV and held-out cohorts.

| **Dataset** | **Site** | **CV ($HC_{train}$)** | **Held-out HC ($HC_{test}$)** |
|---|---|---|---|
| ABIDE I | CALTECH | 50.00 | N/A |
| | CMU | 49.96 | N/A |
| | LEUVEN_1 | 49.96 | N/A |
| | MAX-MUN | 49.87 | 50.00 |
| | NYU | 49.79 | 50.00 |
| | PITT | 49.92 | N/A |
| | SBL | 49.92 | N/A |
| | USM | 49.45 | 50.00 |
| ABIDE II | BNI | 49.74 | 50.00 |
| | IU | 49.87 | N/A |
| ASRB | BRIS | 49.83 | 50.00 |
| | MELB | 47.96 | 50.00 |
| | PERT | 49.45 | N/A |
| | SYDN | 4.91 | 50.00 |
| FEMS | | 49.79 | 50.00 |
| MON | | 38.01 | 50.45 |
| IMPACT | | 44.00 | 51.04 |
| KANMDD | | 49.96 | N/A |
| MITASD | | 50.00 | 50.00 |
| OCDPG | | 49.44 | 50.00 |
| RUSMDD | | 50.00 | 50.00 |
| SPAINOCD | | 45.11 | 49.38 |
| TOP15 | | 40.44 | 52.96 |
| WASHASD | | 49.92 | 50.00 |
| YoDA | | 47.75 | 49.80 |

*N/A = collection sites where data for HCs was < 30, therefore all HC data was included in training set.

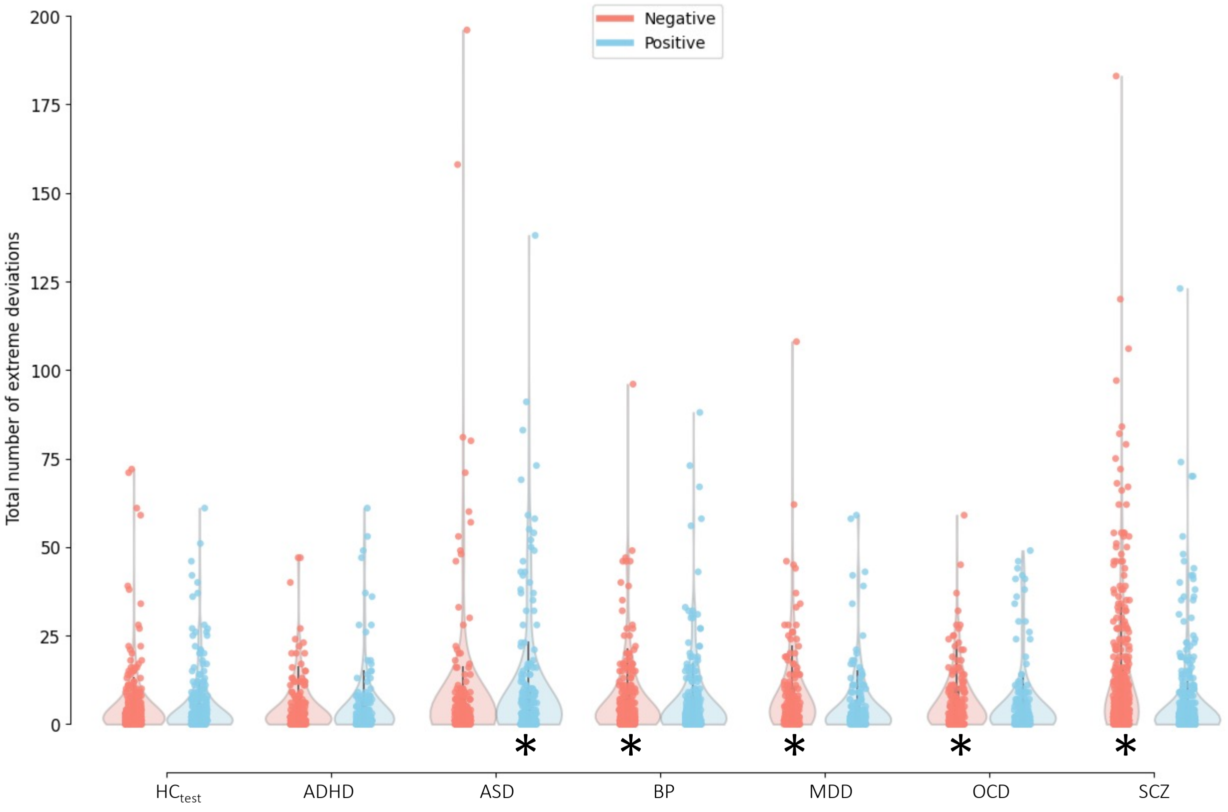


*Figure S4*. **Distribution of person-specific positive (Z > 2.6; blue) and negative (Z < -2.6; red) deviation burden scores (i.e., total number of extreme deviations) in each diagnostic group.** * Indicates clinical groups showing a statistically significant difference in extreme deviation burden compared to the $HC_{test}$ group (Mann Whitney U-test, $p < 0.05$)

*Table S3.* Descriptive statistics for total deviation burden summarised for each disorder.

| | **Negative extreme deviations** | | **Positive extreme deviations** | |
|---|---|---|---|---|
| | % of subjects with at least one extreme deviation | Median deviation burden, median [range] | % of subjects with at least one extreme deviation | Median deviation burden, median [range] |
| $HC_{test}$ | 76.21 | 2 [0 – 72] | 65.43 | 2 [0 – 61] |
| ADHD | 75.82 | 2 [0 – 47] | 69.28 | 1 [0 – 61] |
| ASD | 76.73 | 2 [0 – 196] | 75.74 | 3 [ 0 – 138] |
| BP | 82.46 | 3 [0 – 95] | 71.05 | 2 [0 – 88] |
| MDD | 86.96 | 5 [0 – 108] | 65.22 | 1 [0 – 59] |
| OCD | 79.64 | 4 [0 – 59] | 71.26 | 2 [0 – 49] |
| SCZ | 88.51 | 5 [0 – 183] | 65.27 | 1 [0 – 123] |

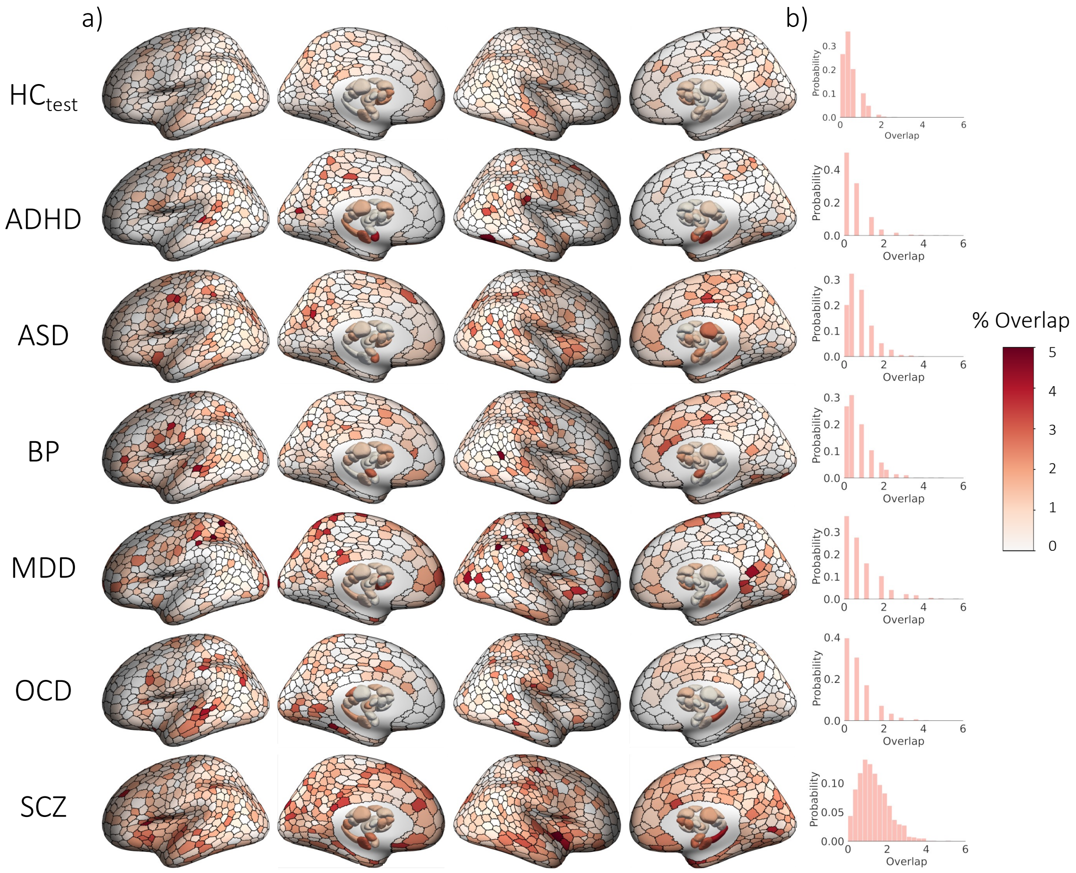


*Figure S5*. **Spatial overlap of extreme negative GMV deviations (Z < - 2.6) in each group.** a) Cortical and subcortical surface renderings showing spatial of overlap in 1032 brain regions, and b) the distribution of overlap percentages observed across all regions.

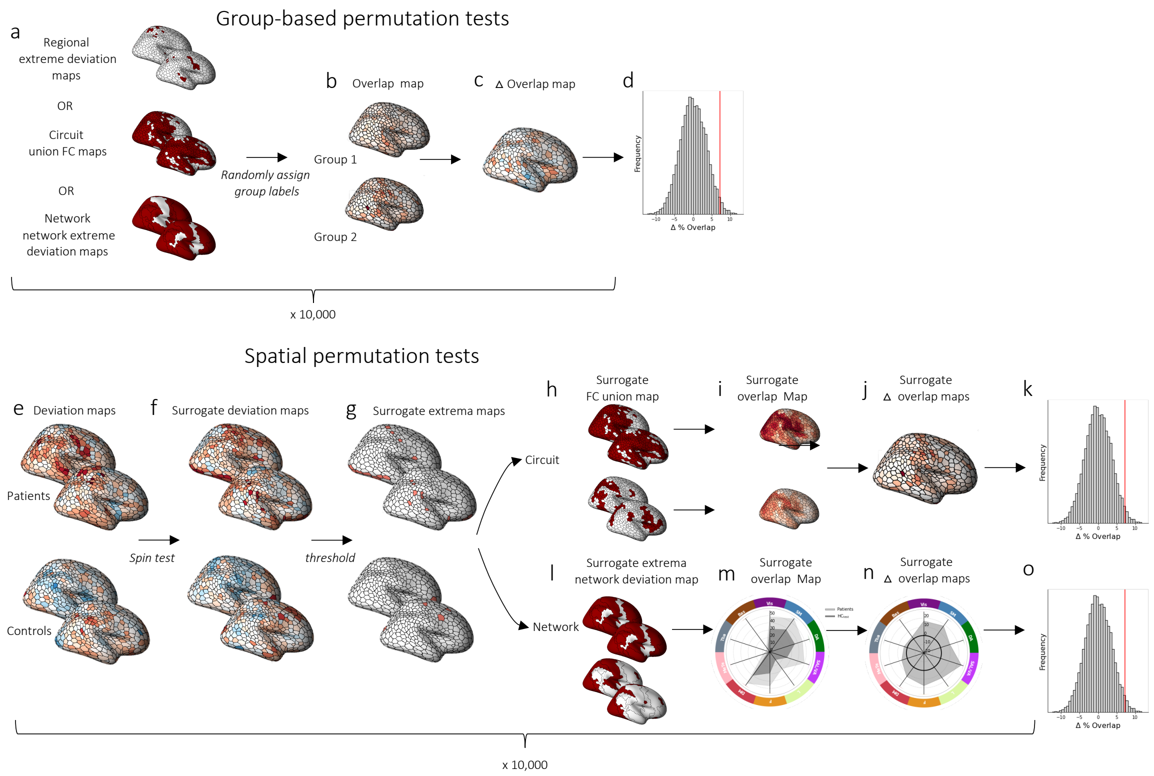


*Figure S6.* **Permutation tests for evaluating region-level, circuit-level, and network-level overlap.** We used two types of permutation test to evaluate different hypotheses. Group-based permutation tests were used to evaluate group differences in region-level, circuit-level, and network-level overlap, regardless of total deviation burden. These tests involved repeating each analysis 10,000 times after shuffling case and control labels. (a) At each iteration, we obtained a new grouping of person-specific deviation maps according to the shuffled group labels. At the regional level, we focused on extreme deviation maps (Fig 2x-x); at the circuit-level, we focused on union FC maps (Fig 3x-x); and at the network level we focused on network extreme deviation maps (Fig 4x-x). (b) For each brain region, we computed an overlap map for the $HC_{test}$ and each clinical group under shuffled group assignment (*Overlap map*). (c) We then subtracted the surrogate $HC_{test}$ overlap map from the surrogate clinical group's overlap map to obtain an overlap difference map *(Δ Overlap map*). Steps (b) and (c) were repeated 10,000 times to derive an empirical distribution of overlap difference maps under the null hypothesis of random group assignment (d). For each brain region, we obtained *p*-values as the proportion of null values that exceeded the observed difference. The second type of permutation test we used was a spatial permutation test. (e) We used the unthresholded deviation maps of each person derived from the normative model to generate an ensemble of surrogate deviation maps for each individual in the test data (f). For cortical regions, the surrogate maps were generated using Hungarian spin tests[19,20]. For subcortical regions, we randomly shuffled deviation values across all subcortical areas (see Methods). (g) We then thresholded the null deviation maps ($Z> |2.6|$) to generate surrogate extreme deviation maps. To evaluate circuit-level group differences in overlap, (h) we obtained individual-specific surrogate FC union maps using the same procedure described in Figure 3a-d. (i) For the $HC_{test}$ group and each clinical group, we calculated surrogate within-group overlap maps. (j) We subtracted the $HC_{test}$ surrogate FC overlap map from each clinical group's surrogate FC overlap map to obtain a surrogate overlap difference map (Δ *Overlap map*). Steps (f) – (j) were repeated 10,000 times to generate (k) a null distribution of circuit-level overlap difference maps for each disorder. To evaluate network-level group differences in overlap, (l) we obtained surrogate network-level extreme deviation maps using the same procedure described in Figure 5a-c. (m) For each clinical group and the control group, we quantified the proportion of individuals showing a surrogate deviation within each network (*Overlap map*). (n) We subtracted the $HC_{test}$ surrogate network overlap map from each clinical group's surrogate overlap (Δ *Overlap map*). Steps (f) – (n) were repeated 10, 000 times to generate (o) a null distribution of overlap difference maps for each disorder. For all tests, statistically significance differences were identified using a threshold of $p_{FDR} < 0.05$, two-tailed.

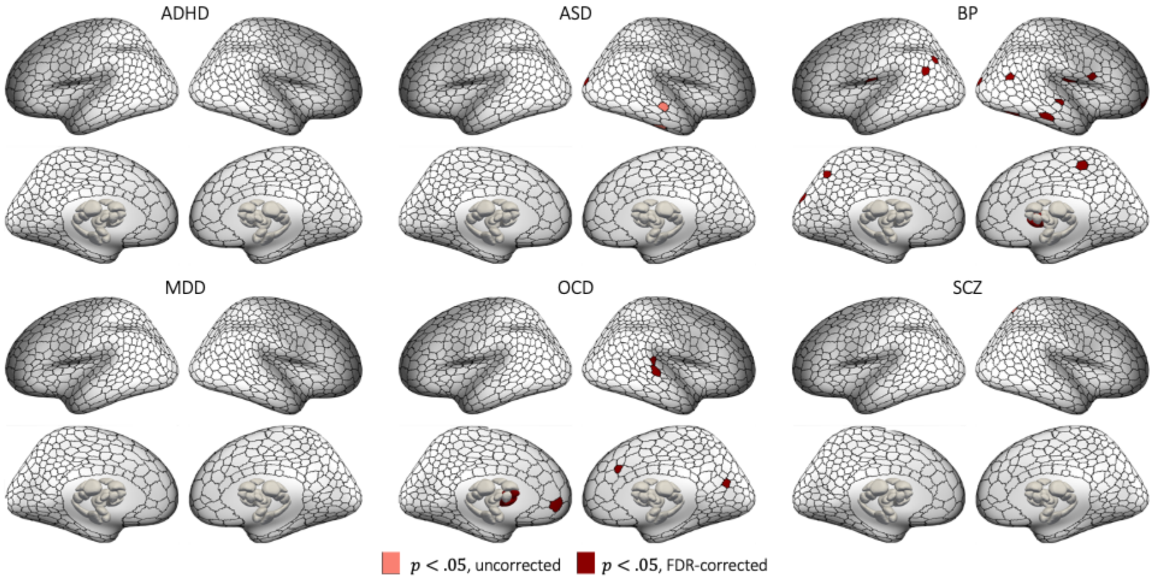


*Figure S7*. **Regions showing greater regional overlap of extreme negative GMV deviations in controls compared to cases.** Statistical maps showing regions with significantly greater overlap in controls, compared to each clinical group in extreme negative deviations ($p < 0.05$, two-tailed, cases<controls).

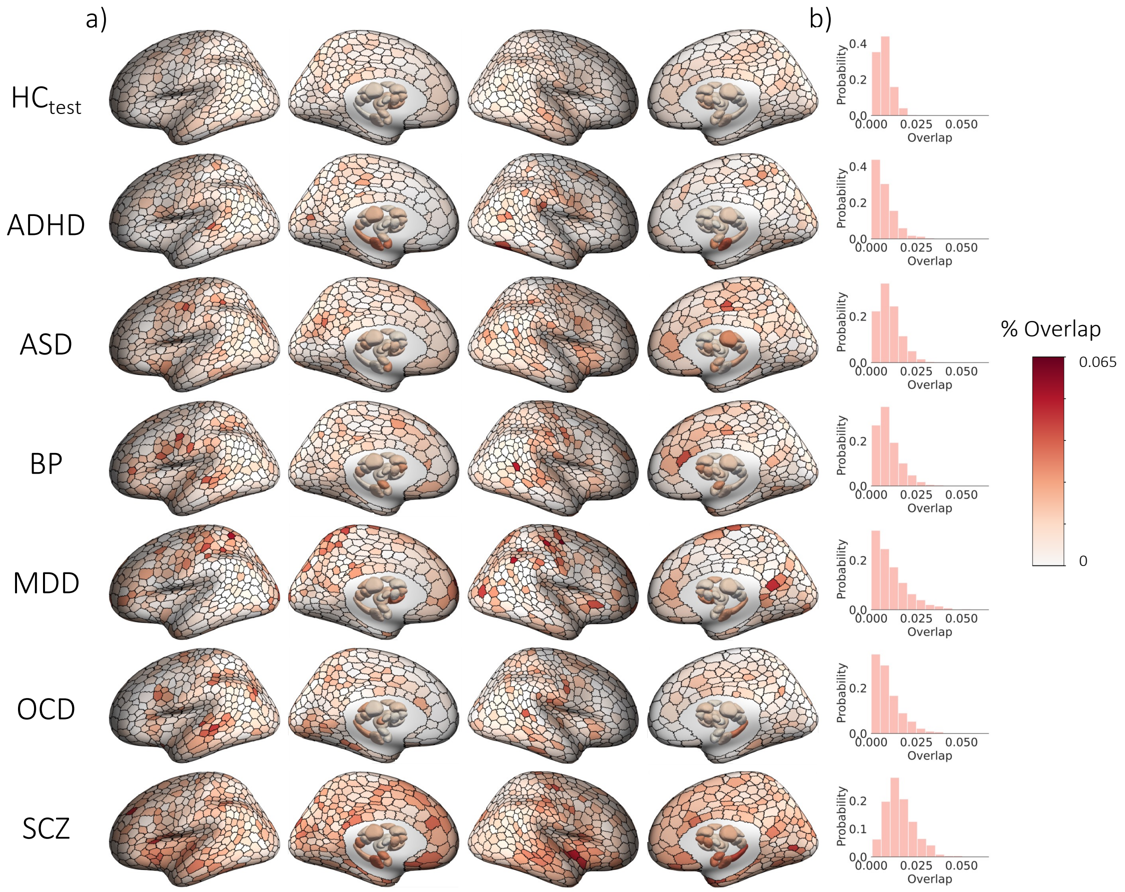


*Figure S8*. **Spatial overlap of extreme negative GMV deviations in each group using a threshold-weighted approach.** a) Cortical and subcortical surface renderings showing spatial of overlap in 1032 brain regions, and b) the distribution of overlap percentages observed across all regions.

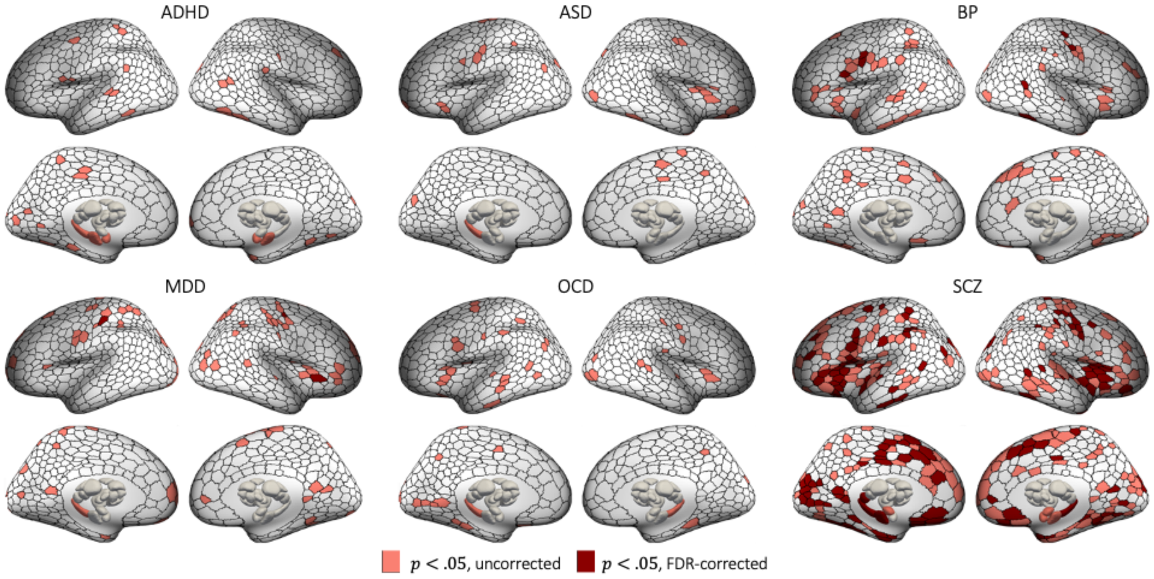


*Figure S9*. **Regions showing greater regional overlap of extreme negative GMV deviations in cases compared to controls, as identified using a weighted-threshold approach.** Statistical maps showing regions with significantly greater overlap in each clinical group, compared to controls in extreme negative deviations (*p* < 0.05, two-tailed, cases>controls).

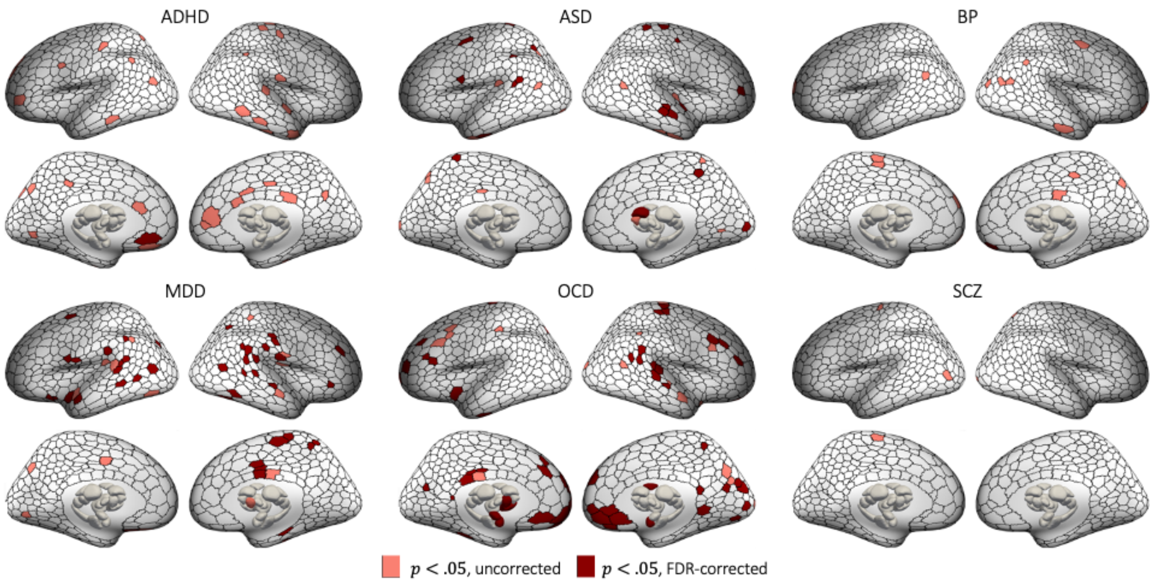


*Figure S10.* **Regions showing greater regional overlap of extreme negative GMV deviations in controls compared to cases, as identified using a weighted-threshold approach.** Statistical maps showing regions with significantly greater overlap in controls, compared to each clinical group in extreme negative deviations ($p < 0.05$, two-tailed, cases<controls).

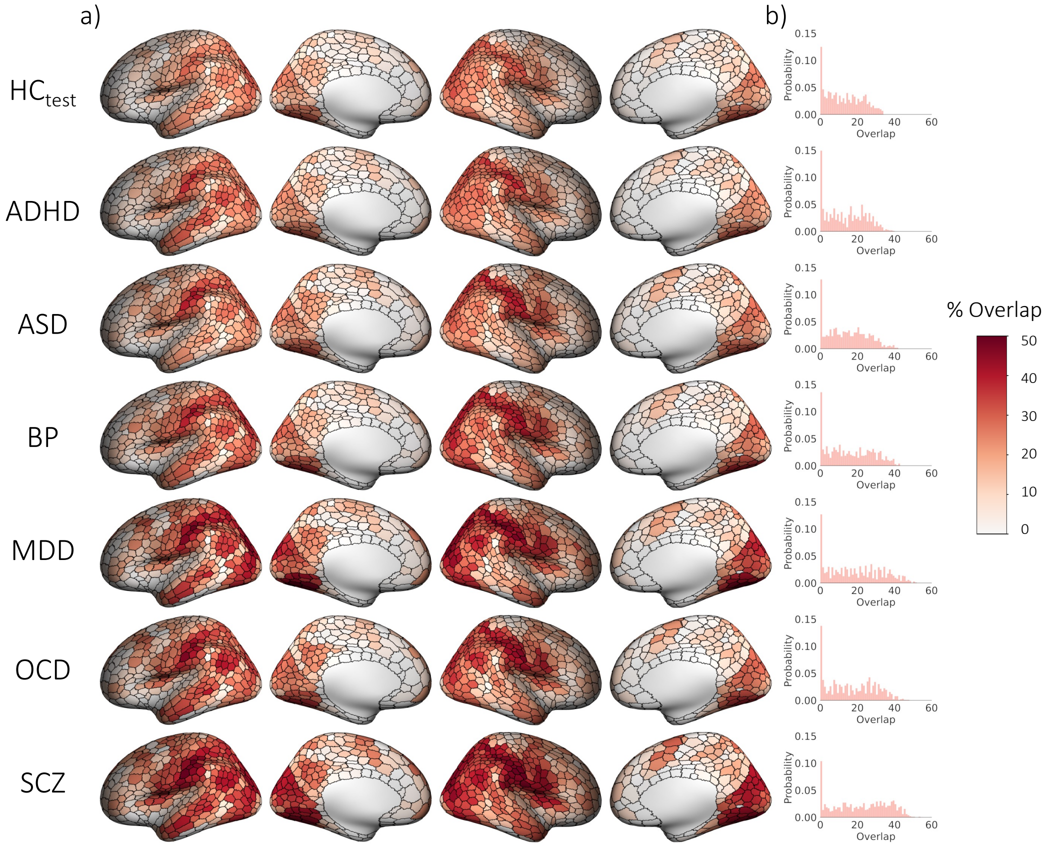


*Figure S11.* **Spatial overlap of regions functionally coupled (vertex-wise threshold $p_{FWE} < 0.025$), to extreme negative deviations (Z < - 2.6) across groups, using a parcel-mapping threshold of 50%).** a)

Cortical surface renderings showing spatial overlap and b) the distribution of overlap percentages observed across all regions.

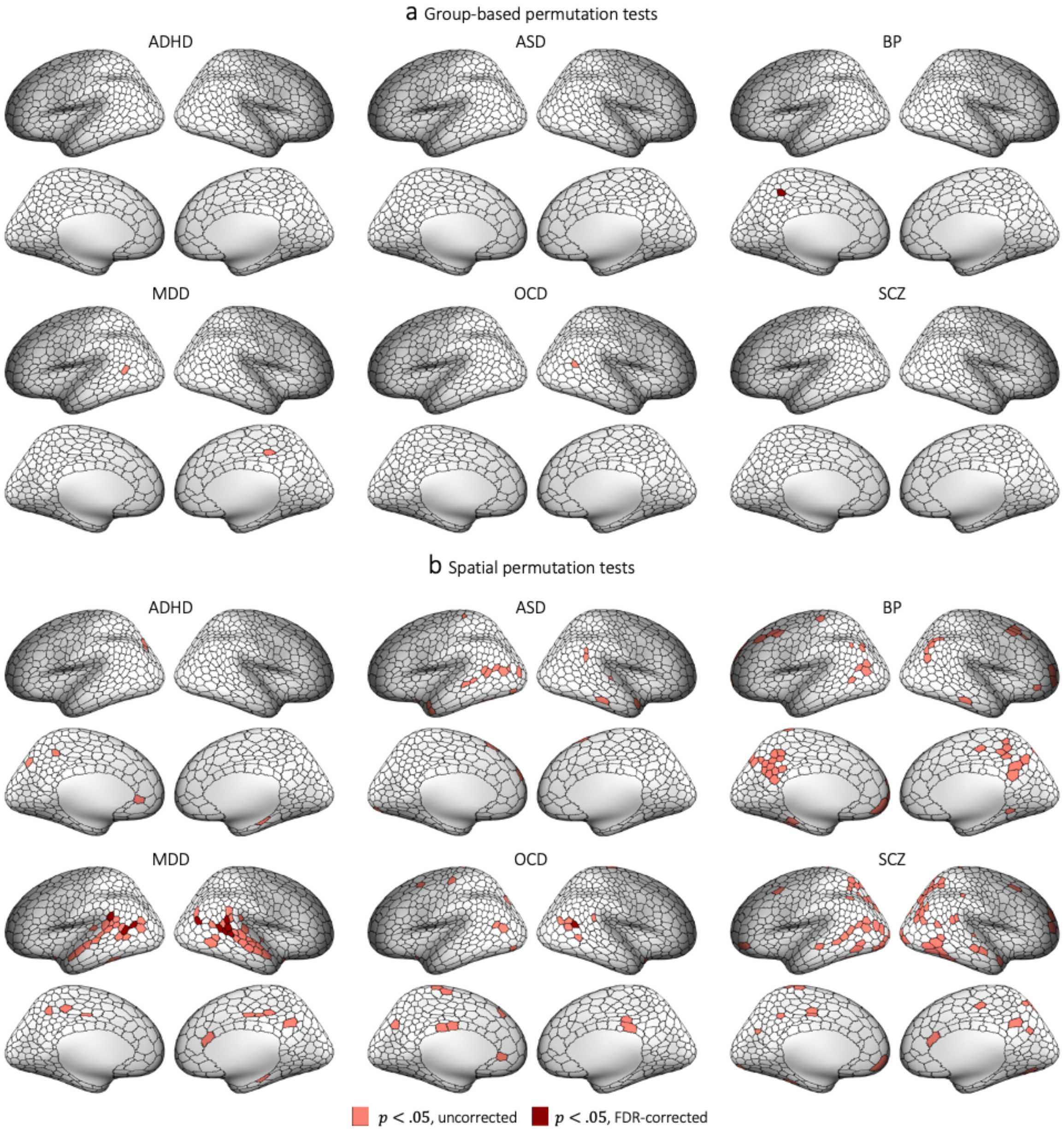


*Figure S12*. **Regions showing greater overlap in areas functionally coupled to extreme negative GMV deviations in controls compared to cases.** Cortical surface renderings showing regions with significantly greater overlap in controls compared to cases in areas functionally coupled to extreme negative deviations ($p < 0.05$, two-tailed, cases<controls). (a) and (b) respectively represent significant areas identified using group-based or spatial permutation tests. The former identifies differences in overlap regardless of group differences in total deviation burden; the latter accounts for these differences and can thus reveal circuits that are preferentially impacted beyond the effects of deviation burden.

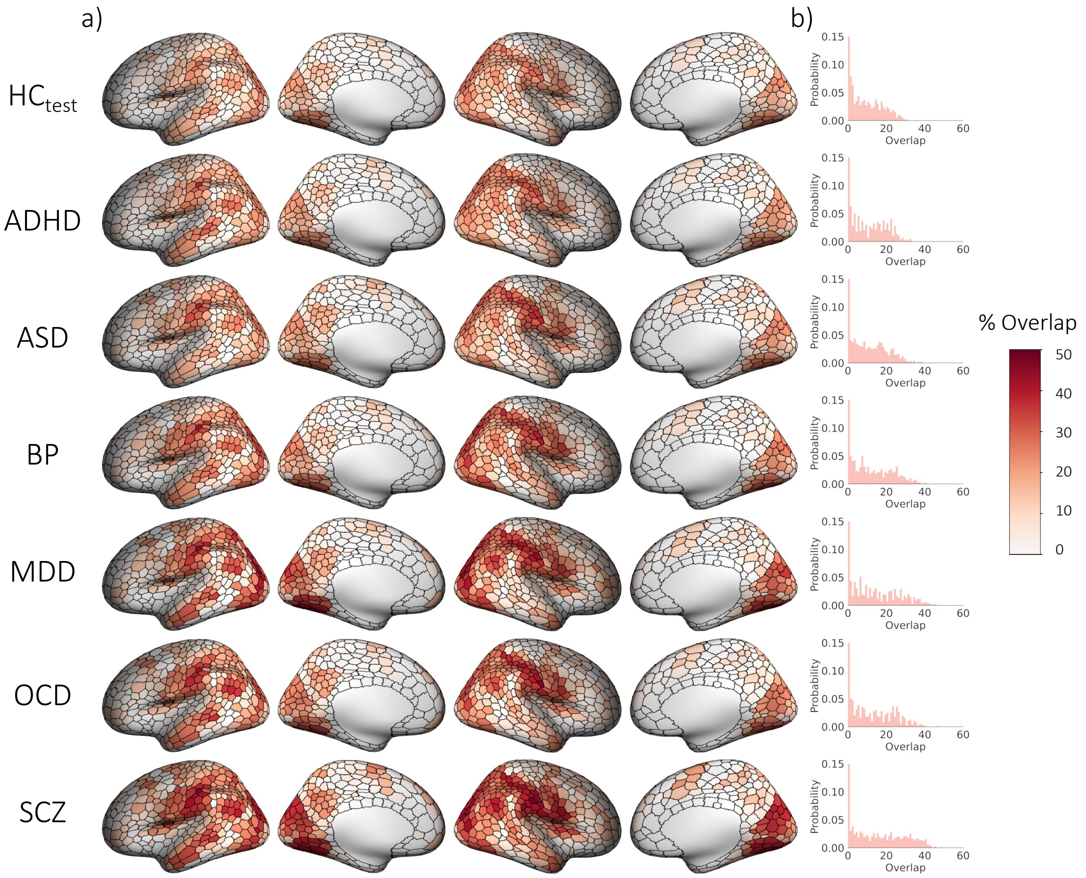


*Figure S13.* **Spatial overlap in regions functionally coupled (vertex-wise threshold $p_{FWE} < 0.025$) to extreme negative deviations (Z < - 2.6) across groups, using a parcel-mapping threshold of 75%.** a) Cortical surface renderings showing spatial of overlap, and b) the distribution of overlap percentages observed across all regions.

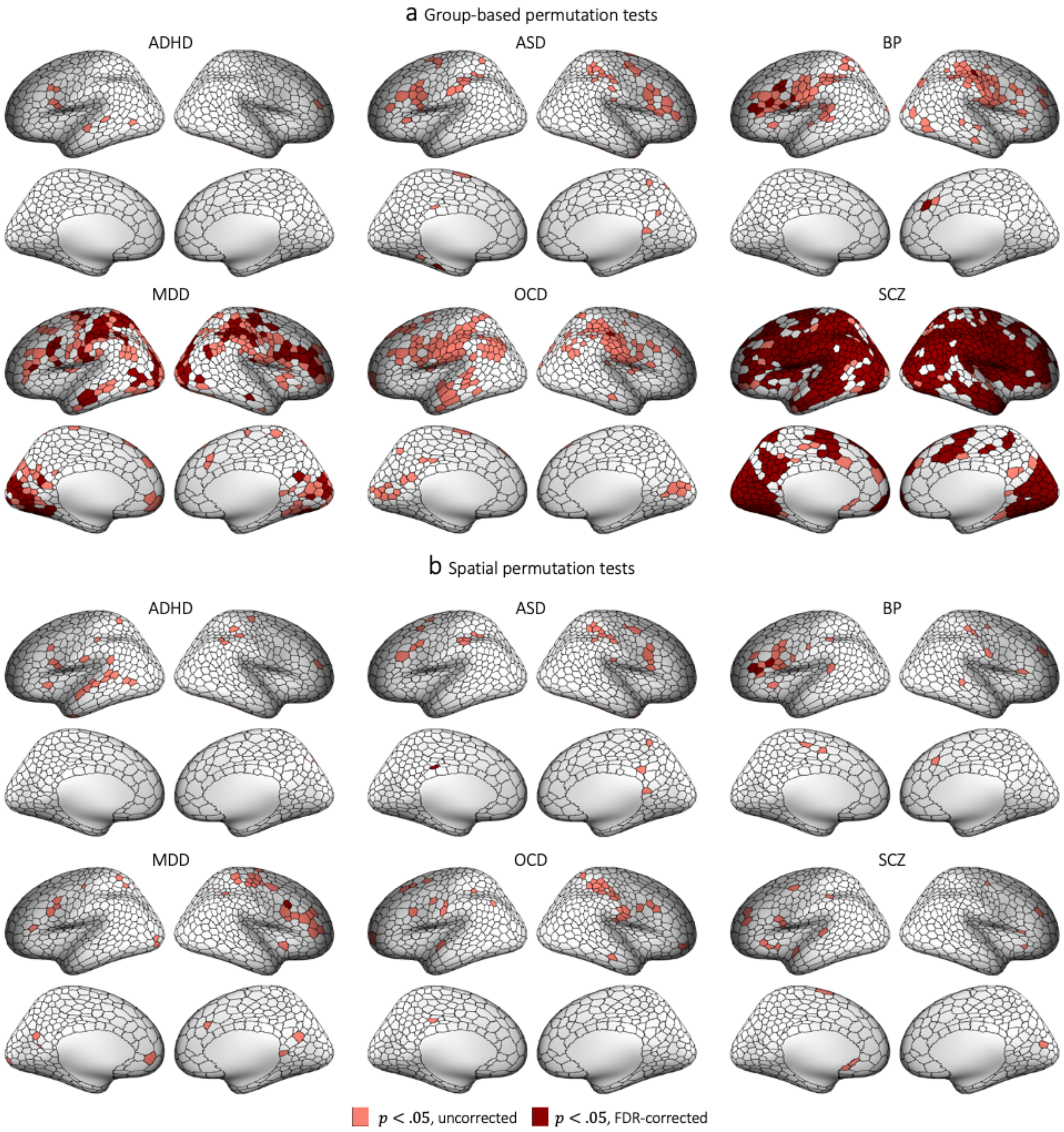


*Figure S14*. **Regions showing greater overlap in areas functionally coupled to extreme negative GMV deviations in cases compared to controls, using parcel-mapping threshold of 75%.** Cortical surface renderings showing regions with significantly greater overlap in cases compared to controls in areas functionally coupled to extreme negative deviations ($p < 0.05$, two-tailed, cases>controls). (a) and (b) respectively represent significant areas identified using group-based or spatial permutation tests. The former identifies differences in overlap regardless of group differences in total deviation burden; the latter accounts for these differences and can thus reveal circuits that are preferentially impacted beyond the effects of deviation burden.

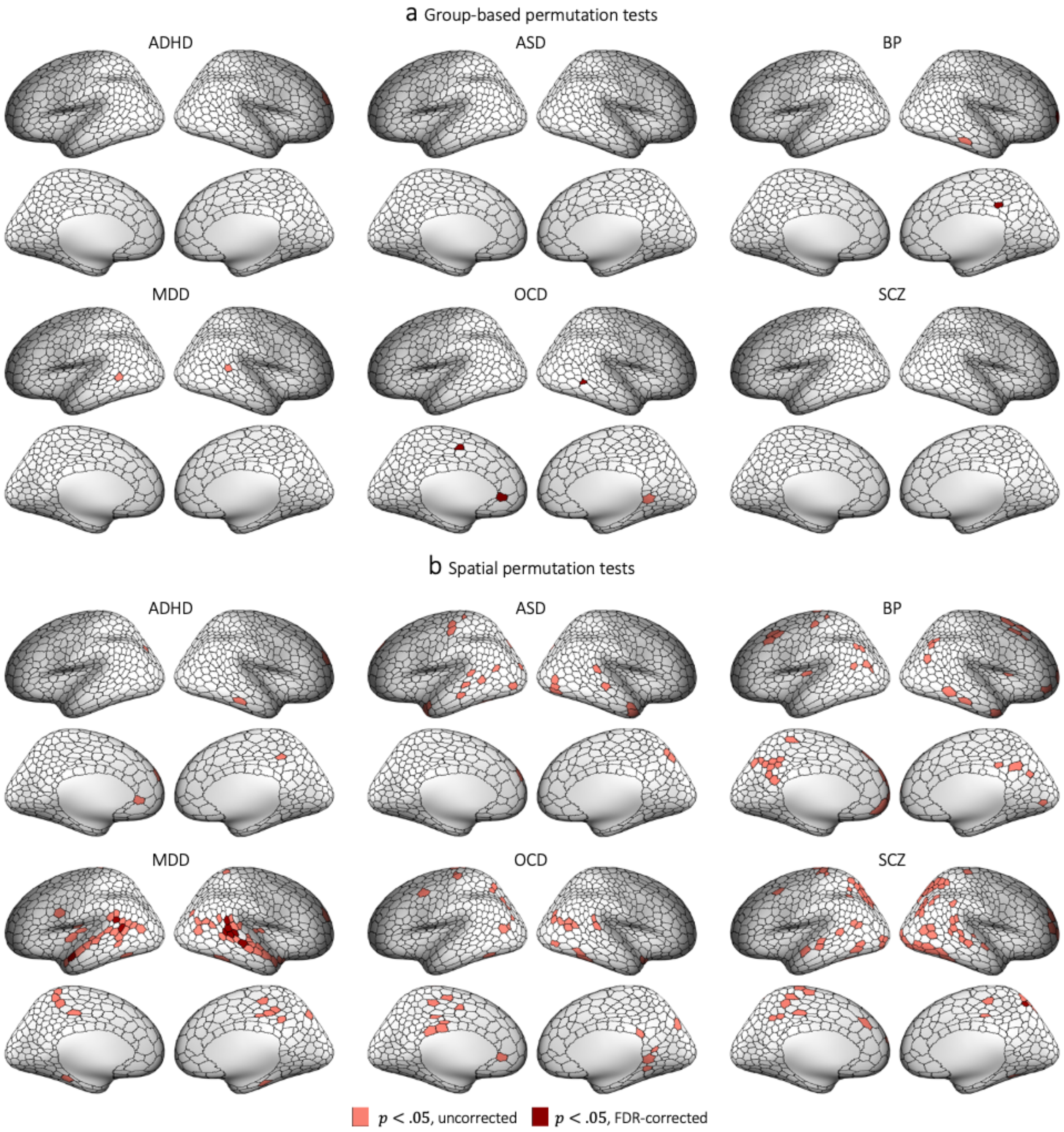


*Figure S15*. **Regions showing greater overlap in areas functionally coupled to extreme negative GMV deviations in controls compared to cases, using parcel-mapping threshold of 75%.** Cortical surface renderings showing regions with significantly greater overlap in controls compared to cases in areas functionally coupled to extreme negative deviations ($p < 0.05$, two-tailed, cases<controls). (a) and (b) respectively represent significant areas identified using group-based or spatial permutation tests. The former identifies differences in overlap regardless of group differences in total deviation burden; the latter accounts for these differences and can thus reveal circuits that are preferentially impacted beyond the effects of deviation burden.

*Table S4.* **The degree of spatial overlap (%) in each network for each group.**

| | Vis | SM | DA | SAL/VA | L | F | DM | MeTe | Tha | Bas |
|---|---|---|---|---|---|---|---|---|---|---|
| $HC_{test}$ | 26.77 | 37.18 | 24.91 | 20.82 | 13.38 | 24.91 | 35.69 | 2.60 | 1.86 | 4.83 |
| ADHD | 29.41 | 40.52 | 39.22 | 30.72 | 11.11 | 34.64 | 36.60 | 10.46 | 2.61 | 3.27 |
| ASD | 34.16 | 40.10 | 39.60 | 32.67 | 14.85 | 36.14 | 35.64 | 5.45 | 4.46 | 4.46 |
| BP | 33.77 | 49.12 | 42.54 | 33.33 | 17.54 | 40.35 | 47.81 | 3.95 | 2.19 | 4.82 |
| MDD | 50.93 | 49.69 | 48.45 | 35.40 | 24.84 | 43.48 | 54.66 | 5.59 | 3.73 | 7.45 |
| OCD | 40.12 | 48.50 | 37.13 | 38.32 | 16.77 | 34.73 | 47.31 | 2.99 | 2.40 | 4.79 |
| SCZ | 48.30 | 56.14 | 47.00 | 49.61 | 28.20 | 46.48 | 53.26 | 9.66 | 4.96 | 7.83 |

| | |
|---|---|
| *VIS* | *Visual* |
| *SM* | *Somatomotor* |
| *DA* | *Dorsal attention* |
| *SAL/VA* | *Salience/ventral attention* |
| *L* | *Limbic* |
| *F* | *Frontoparietal* |
| *DM* | *Default mode* |
| *MeTe* | *Medial Temporal* |
| *Tha* | *Thalamus* |
| *Bas* | *Basal Ganglia* |

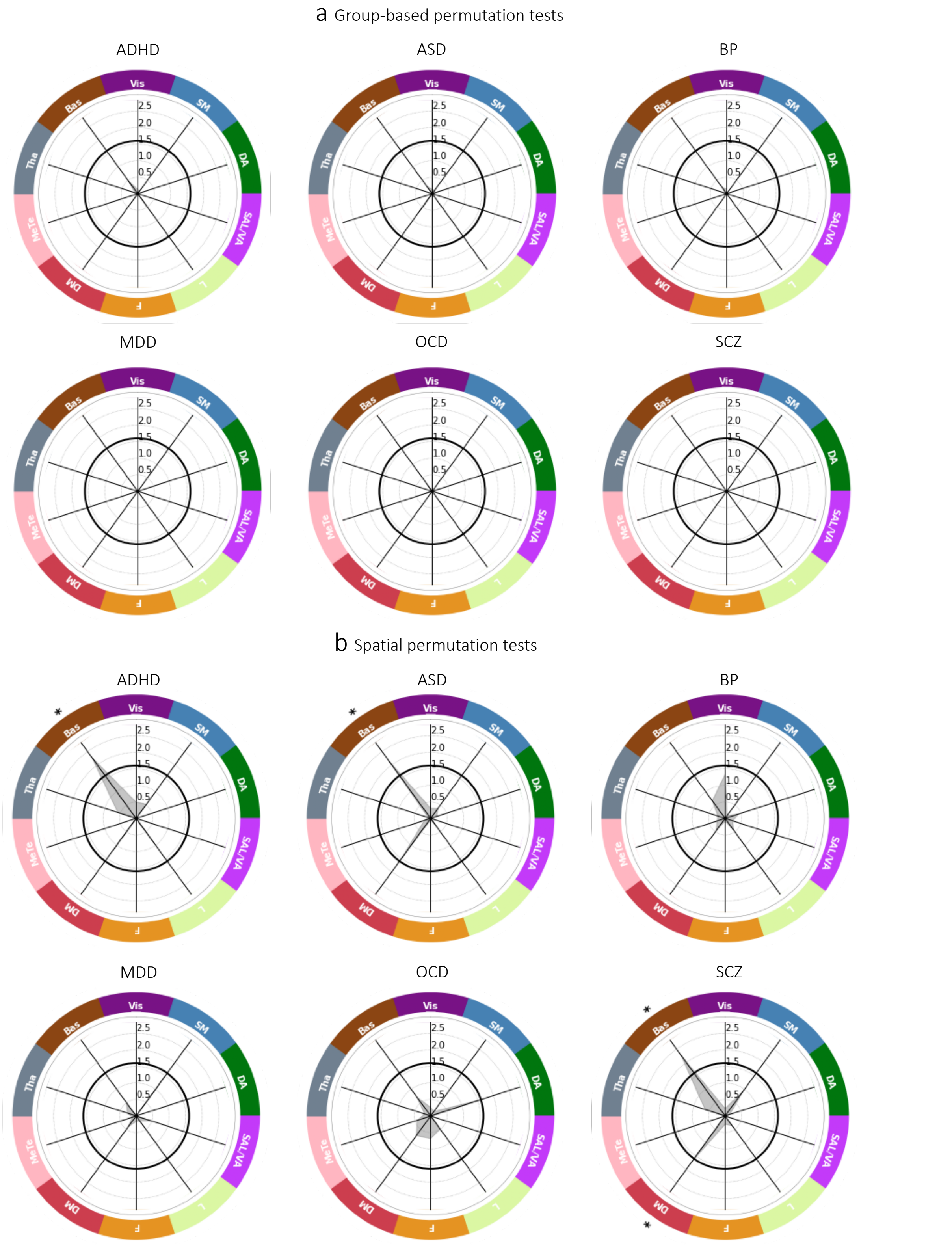


*Figure S16*. **Functional networks showing greater overlap in extreme negative GMV deviations in controls compared to cases.** The network-level -$\log_{10}$ p-values associated with difference in percent overlap for extreme negative GMV deviations between each clinical group and the $HC_{test}$ cohort. ** corresponds to $p_{FDR} < 0.05$, two-tailed, cases<controls, * corresponds to $p_{uncorrected} < 0.05$, two-tailed, cases<controls. The solid black line

indicates $-\log_{10} p = 1.6$ ($p$=0.05, two-tailed, uncorrected). (f) and (g) identify networks showing significant differences under group-based or spatial permutation testing, respectively.

### Analysis of positive GMV deviations

The analyses presented in the main text focus on understanding extreme negative GMV deviations, representing areas where volume is lower than normative expectations, given that GMV reductions are often emphasized in the psychiatric neuroimaging literature. For completeness, we repeated the same analyses for positive GMV extreme deviations, representing areas where volume was higher than normative expectations. At the regional level, the overlap in the location of extreme deviations never exceeded 6% (ADHD: 5.23%, ASD: 3.96%, BP: 4.82%, MDD: 5.59%, OCD: 4.19%, SCZ: 5.22% HC: 2.60%) and there were very few regions showing significant case-control differences in overlap (Figure S16-18). Overlap was higher at the circuit level, with a maximum of 40% across all regions and disorders (Figure S19). Group-based permutation testing revealed significantly greater circuit-level overlap in individuals diagnosed with ASD compared to controls in ~20% of regions, which were predominately located in visual, parietal, and frontal cortices. No other differences survived FDR correction (Figure S20a-21a). Similarly, spatial permutation testing only identified isolated areas in pregenual cingulate and right lateral prefrontal cortex as showing significantly greater overlap in MDD (Figure S20b-21b). At the network level, observed group overlaps were as high as 48% (Table S4), with group-based permutation testing identifying significantly greater overlap in all cortical networks except the default mode network in ASD and the basal ganglia in SCZ ($p < 0.05$, two-tailed), compared to controls (Figure S22a). However, only the latter difference was also observed with spatial permutation tests, being accompanied by additional evidence of greater overlap in the salience/ventral attention network in SCZ (Figure S22b). The medial temporal lobe showed significantly greater overlap in controls compared to individuals diagnosed with SCZ (Figure S23).

Taken together, these findings indicate that positive GMV deviations are more heterogeneous than negative deviations. Elevated circuit-level and network-level overlap was particularly prominent in ASD under group-based permutation testing, and implicated areas of medial and lateral parietal, temporal and prefrontal cortex at the circuit level and all cortical systems except the default mode network. These differences were not apparent with spatial permutation testing, indicating that they were largely driven by the elevated positive GMV burden of individuals diagnosed with ASD (Figure S21). ASD has been associated with dysregulated and accelerated brain growth, particularly in the temporal, parietal, and frontal association cortices during early childhood[21], although whether these increases persist into adulthood has been unclear.

Differences in circuit-level and network-level overlap for positive GMV deviations in other disorders were less pronounced. In MDD, the greater circuit-level overlap in left pregenual cingulate and anterior right lateral PFC is consistent with the known roles of these areas in regulating emotion[22] and cognitive control[23], respectively. The greater overlap observed in the basal ganglia of people with SCZ may be attributable to the effects of antipsychotics, which can cause volumetric expansion in this region[24].

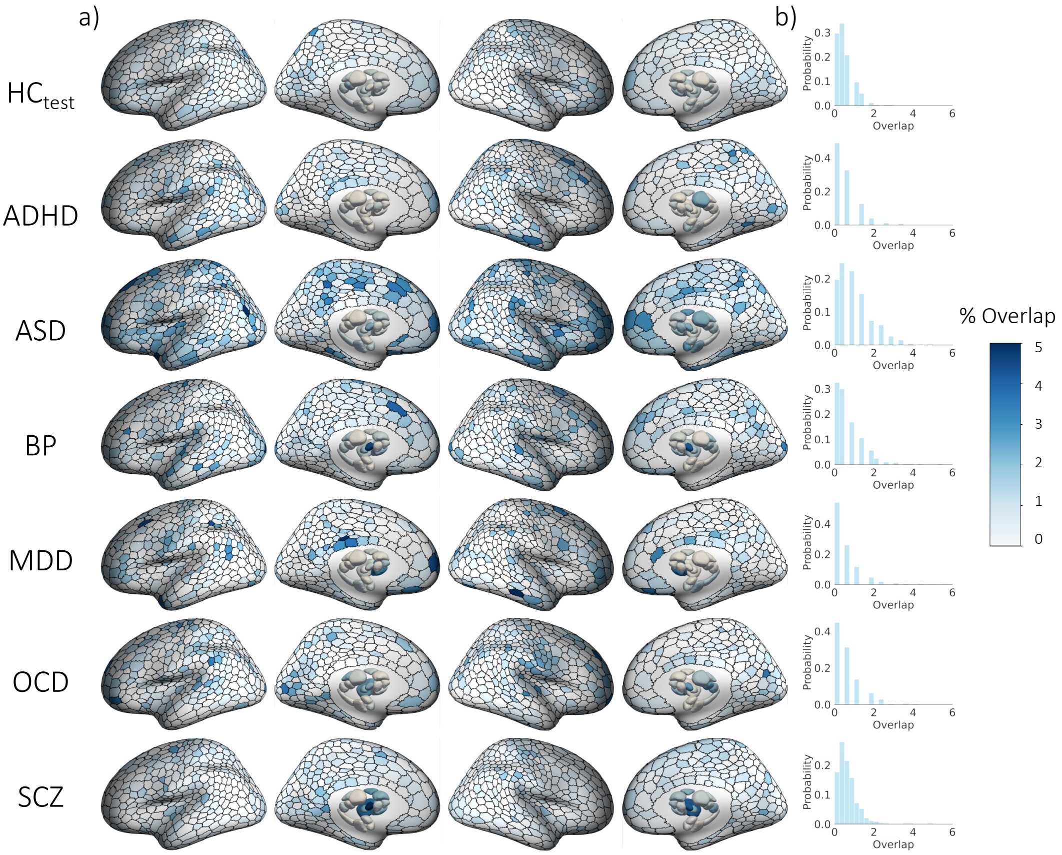


*Figure S17*. **Spatial overlap of extreme positive GMV deviations (Z > 2.6) in each group.** a) Cortical and subcortical surface renderings showing spatial of overlap in 1032 brain regions, and b) the distribution of overlap percentages observed across all regions.

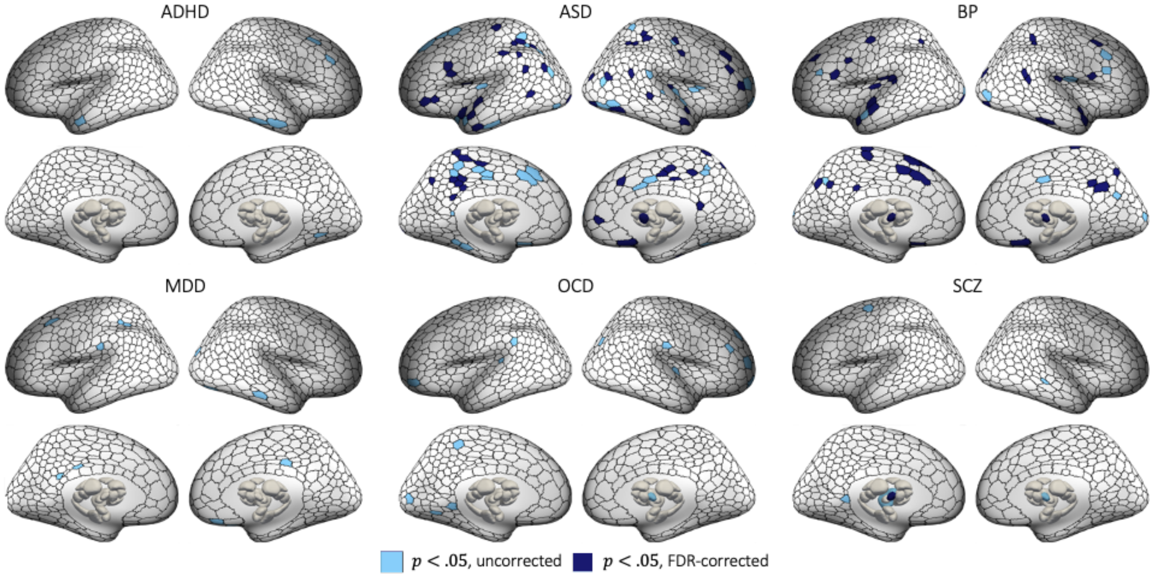


*Figure S18*. **Regional heterogeneity of extreme positive GMV deviations in each disorder.** Cortical and subcortical surface renderings showing regions with significantly greater overlap of extreme positive GMV deviations in cases compared to controls.

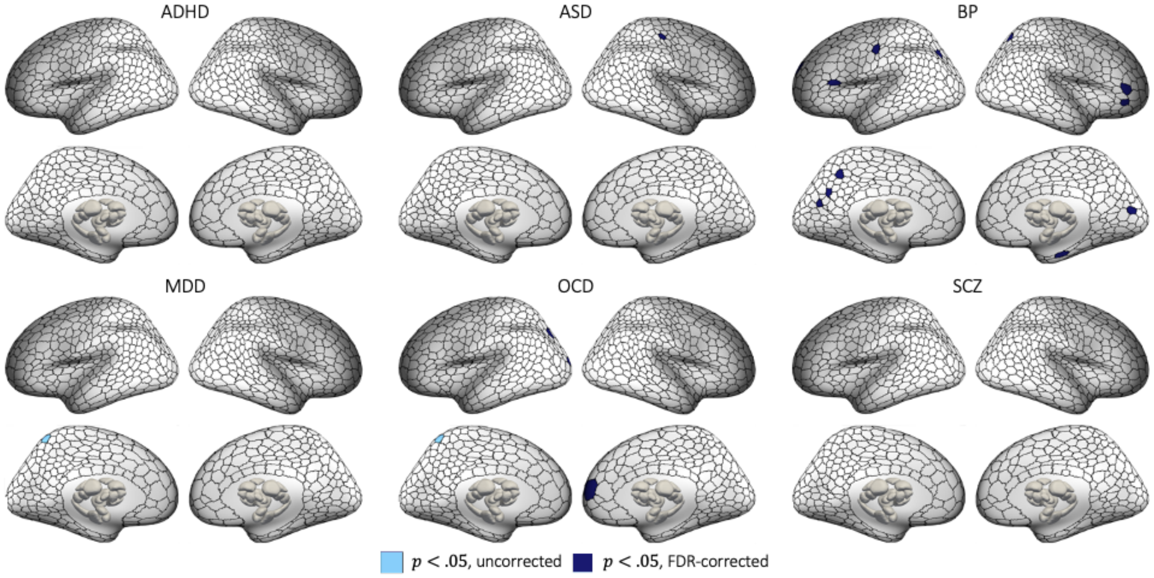


*Figure S19*. **Regions showing greater regional overlap of extreme positive GMV deviations in controls compared to cases.** Statistical maps showing regions with significantly greater overlap in controls, compared to cases in extreme positive deviations ($p < 0.05$, two-tailed, cases<controls).

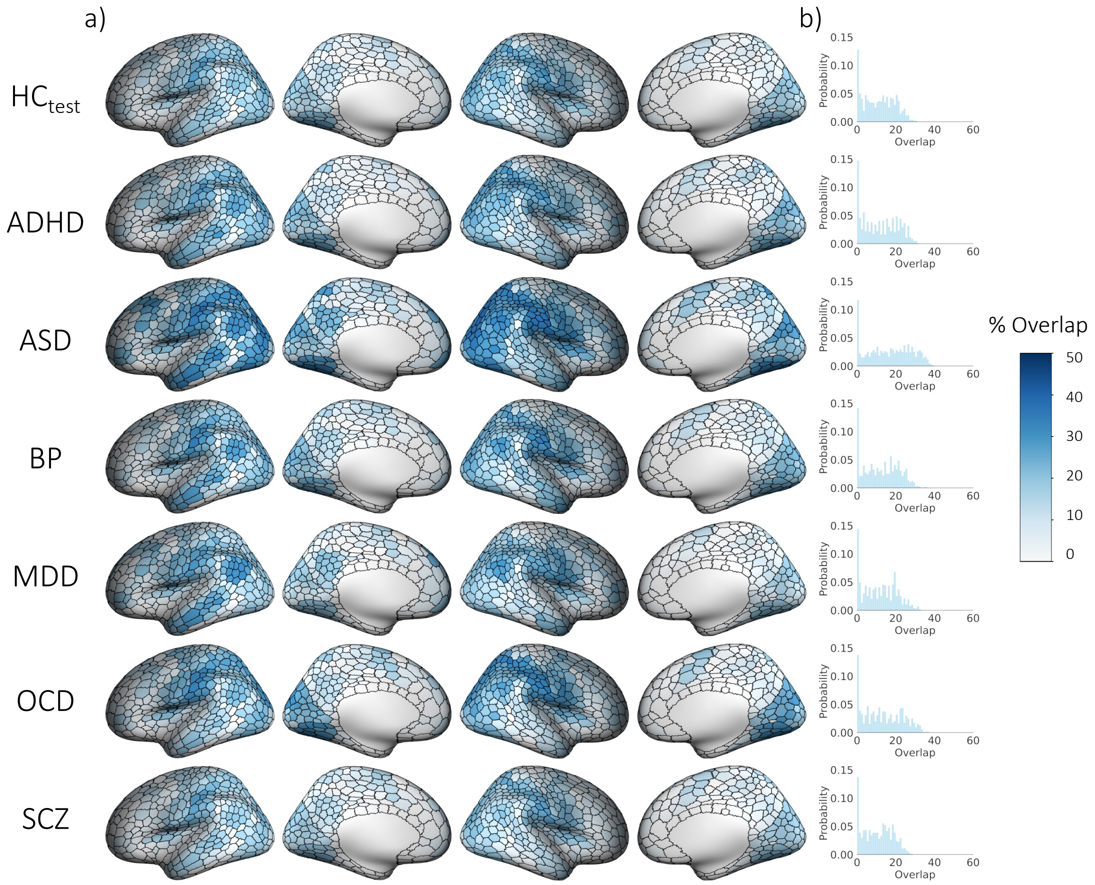


*Figure S20.* **Spatial overlap in regions functionally coupled (vertex-wise threshold $p_{FWE} < 0.025$), to extreme positive deviations (Z > 2.6) across groups, using a parcel-mapping threshold of 50%).** a) Cortical surface renderings showing spatial overlap, and b) the distribution of overlap percentages observed across all regions.

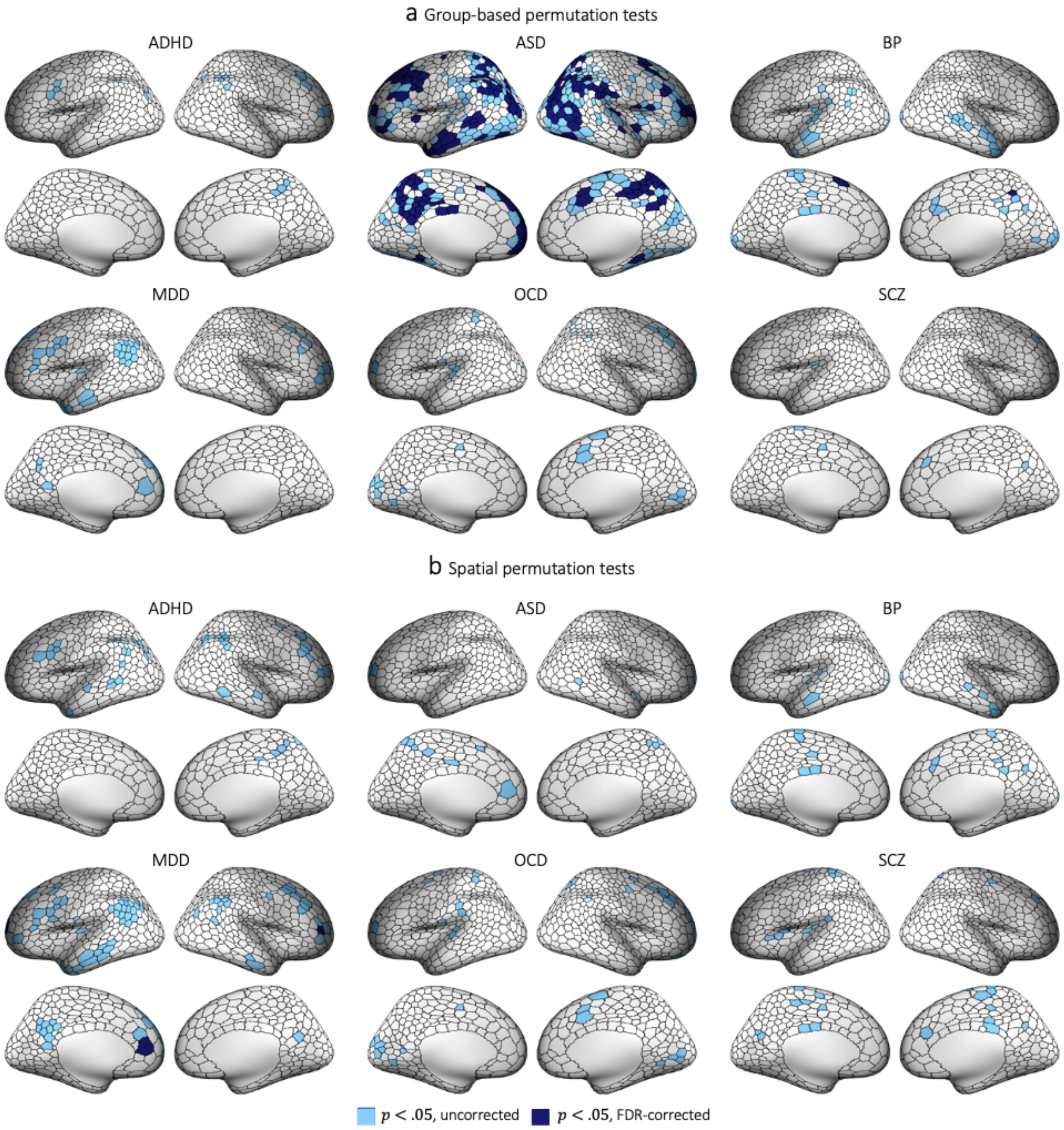


*Figure S21.* **Regions showing greater overlap in areas functionally coupled to extreme positive GMV deviations in cases compared to controls.** Group differences in circuit-level overlap were evaluated with respect to two empirical null models (see Figure S6 for details). (a) and (b) show **c**ortical surface renderings of regions with significantly greater overlap in cases compared to controls in areas functionally coupled to extreme deviations identified using group-based or spatial permutation tests, respectively.

*Table S5.* **The degree of spatial overlap (%) in each network for each group.**

| | Vis | SM | DA | SAL/VA | L | F | DM | MeTe | Tha | Bas |
|---|---|---|---|---|---|---|---|---|---|---|
| HC | 26.77 | 37.17 | 24.91 | 20.82 | 13.38 | 24.91 | 35.69 | 2.60 | 1.86 | 4.83 |
| ADHD | 27.45 | 30.07 | 30.72 | 19.61 | 10.46 | 32.03 | 33.33 | 2.61 | 2.61 | 2.61 |
| ASD | 37.13 | 48.02 | 41.58 | 39.11 | 25.25 | 39.60 | 45.05 | 5.94 | 4.46 | 7.43 |
| BP | 33.33 | 46.49 | 27.63 | 27.63 | 16.23 | 28.07 | 35.09 | 0.88 | 3.51 | 11.40 |
| MDD | 22.98 | 32.30 | 21.74 | 19.25 | 18.01 | 34.16 | 42.86 | 2.48 | 0.00 | 5.59 |
| OCD | 35.93 | 40.12 | 30.54 | 23.95 | 13.77 | 27.54 | 34.73 | 1.20 | 2.99 | 7.78 |
| SCZ | 24.80 | 36.29 | 24.02 | 25.59 | 11.75 | 22.72 | 31.07 | 0.26 | 3.13 | 18.80 |

| | |
|---|---|
| *VIS* | *Visual* |
| *SM* | *Somatomotor* |
| *DA* | *Dorsal attention* |
| *SAL/VA* | *Salience/ventral attention* |
| *L* | *Limbic* |
| *F* | *Frontoparietal* |
| *DM* | *Default mode* |
| *MeTe* | *Medial Temporal* |
| *Tha* | *Thalamus* |
| *Bas* | *Basal Ganglia* |

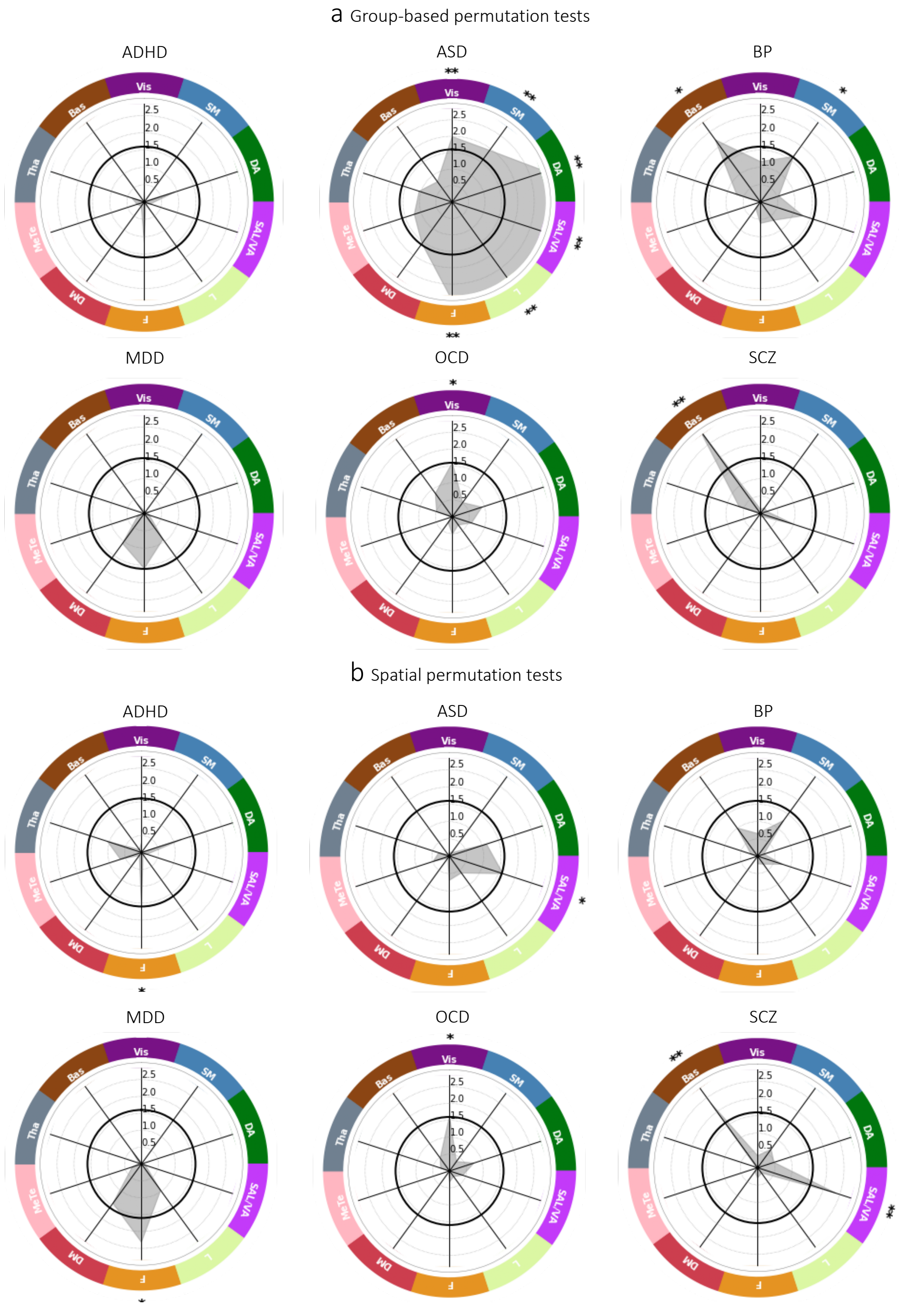


*Figure S23*. **Functional networks showing greater overlap in extreme positive GMV deviations in cases compared to controls.** The network-level -$\log_{10}$ p-values associated with difference in percent overlap for extreme positive GMV deviations between each clinical group and the $HC_{test}$ cohort. ** corresponds to $p_{FDR}$ < 0.05, two-tailed, cases>controls, * corresponds to $p_{uncorrected}$ < 0.05, two-tailed, cases>controls. The solid black line indicates -$\log_{10}$ p = 1.6 ($p$=0.05, two-tailed, uncorrected). (a) and (b) identify networks showing significant differences under group-based or spatial permutation testing, respectively.

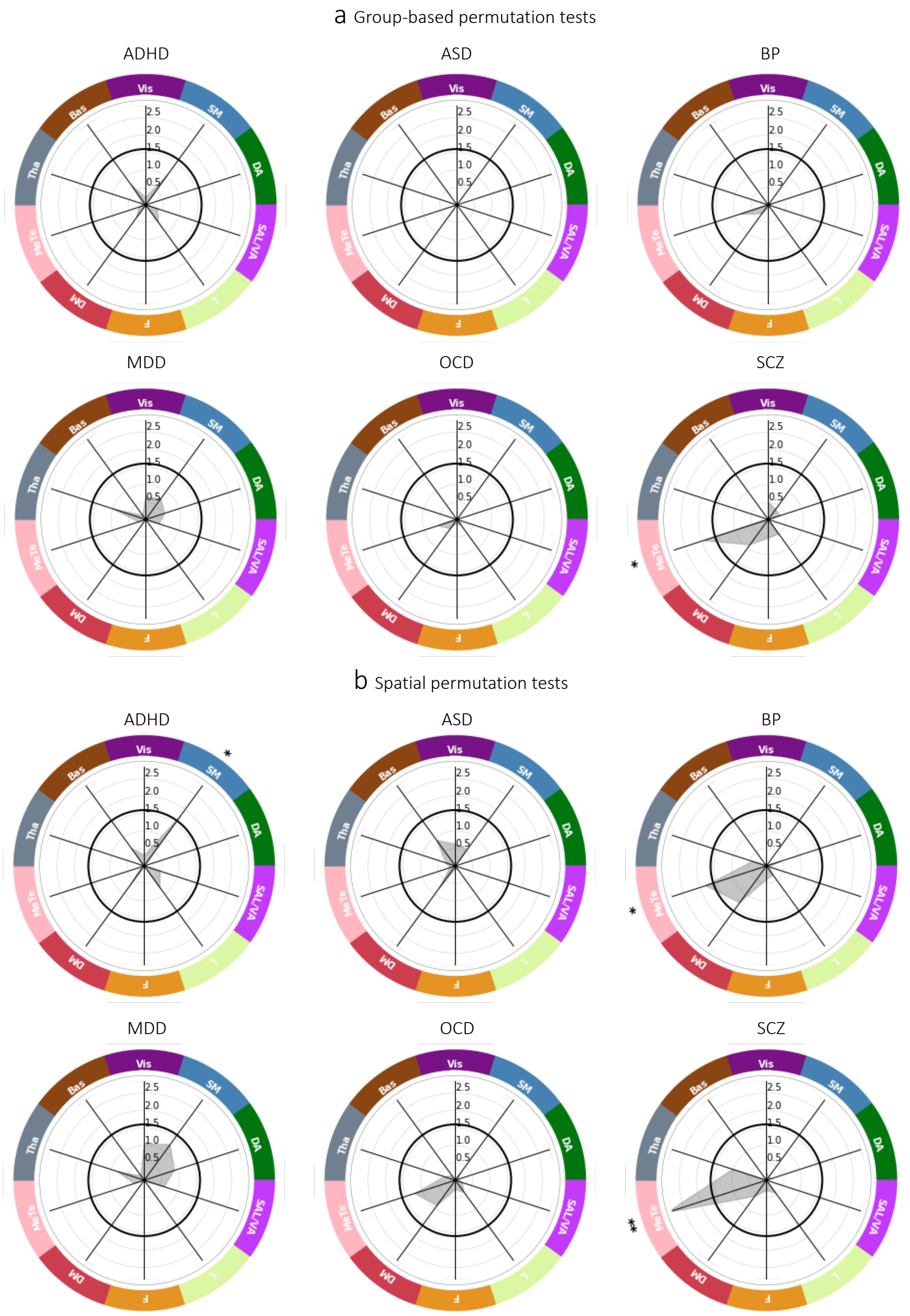


*Figure S24.* **Functional networks showing greater overlap in extreme positive GMV deviations in controls compared to cases.** The network-level $-\log_{10}$ p-values associated with difference in percent overlap for extreme positive GMV deviations between each clinical group and the $HC_{test}$ cohort. ** corresponds to $p_{FDR} < 0.05$, two-tailed, cases<controls, * corresponds to $p_{uncorrected} < 0.05$, two-tailed, cases<controls. The solid black line indicates $-\log_{10}$ p = 1.6 ($p$=0.05, two-tailed, uncorrected). (a) and (b) identify networks showing significant differences under group-based or spatial permutation testing, respectively.
